# Quantification of measurable residual disease using duplex sequencing in adults with acute myeloid leukemia

**DOI:** 10.1101/2023.03.26.23287367

**Authors:** Laura W. Dillon, Jake Higgins, Hassan Nasif, Megan Othus, Lan Beppu, Thomas H. Smith, Elizabeth Schmidt, Charles C. Valentine, Jesse J. Salk, Brent L Wood, Harry P. Erba, Jerald P. Radich, Christopher S. Hourigan

## Abstract

The presence of measurable residual disease (MRD) is strongly associated with treatment outcomes in acute myeloid leukemia (AML). Despite the correlation with clinical outcomes, MRD assessment has yet to be standardized or routinely incorporated into clinical trials. Discrepancies have been observed between different techniques for MRD assessment and there remains a need to compare centralized, high-quality multiparametric flow cytometry (MFC) and ultrasensitive next-generation sequencing (NGS) in AML patients with diverse mutational profiles. In 62 patients with AML, aged 18-60, in first complete remission after intensive induction therapy on the randomized phase 3 SWOG-S0106 clinical trial, MRD detection by MFC was compared with a 29 gene panel utilizing duplex sequencing (DS), an NGS method that generates double-stranded consensus sequences to reduce false positive errors. Using DS, detection of a persistent mutation utilizing defined criteria was seen in 22 (35%) patients and was strongly associated with higher rates of relapse (68% vs 13% at year 5; HR, 8.8; 95% CI, 3.2-24.5; P<0.001) and decreased survival (32% vs 82% at year 5; HR, 5.6; 95% CI, 2.3-13.8; P<0.001). MRD as defined by DS strongly outperformed MFC, which was observed in 10 (16%) patients and marginally associated with higher rates of relapse (50% vs 30% at year 5; HR, 2.4; 95% CI, 0.9-6.7; P=0.087) and decreased survival (40% vs 68% at year 5; HR, 2.5; 95% CI, 1.0-6.3; P=0.059). Furthermore, the prognostic significance of DS MRD status at the time of remission was similar on both randomized arms of the trial, predicting S0106 clinical trial outcomes. These findings suggest that DS is a powerful tool that could be used in patient management and for early treatment assessment in clinical trials.

## Introduction

Acute myeloid leukemia (AML) is a rare blood cancer diagnosed in approximately 20,000 Americans annually. While most patients treated with chemotherapy will achieve an initial complete remission, less than one-third are expected to survive after five years.^1, 2^

Measurable residual disease (MRD) is the presence of leukemia below the threshold set for remission by traditional clinical criteria but detectable with higher sensitivity approaches.^3^ The presence of MRD is strongly associated with treatment outcomes.^4, 5^ However, despite being well established as correlated with the antileukemic effect of treatment interventions^6-11^, clinical implementation has been limited. While no standard technique is currently used for AML MRD testing^12^, multiple methodologies exist including detection of aberrant cell surface protein expression by multiparametric flow cytometry (MFC) or detection of genetic alterations by molecular assays^13^.

MFC has been widely used for AML MRD detection, but there are concerns that inter-laboratory variability and a lack of standardization could limit applicability of the technique on a broader scale.^14, 15^ MFC and next-generation sequencing (NGS) have been found to provide discordant MRD results^16^, potentially capturing different residual cell populations. Furthermore, while MRD detection of certain highly prevalent genetic variants, including *FLT3* internal tandem duplications (*FLT3*-ITD) and *NPM1* insertions, by next-generation sequencing (NGS) has been shown to be strongly correlated with adverse clinical outcomes^10, 11, 17^, decentralized flow cytometry on the same patients was not prognostic.^11^ There remains a need to compare AML MRD assessment using both centralized, high-quality MFC and ultra-sensitive NGS for detection of a broad range of variants in the same patients.

The SWOG Cancer Research Network S0106 study was an open-label randomized phase 3 clinical trial of adults aged 18-60 with previously untreated *de novo* non-APL AML comparing standard induction therapy with daunorubicin (60 mg/m2 IV D1,2,3) and cytarabine (100mg/m^2^/d CI D1-7) (“DA”) against the combination of daunorubicin (45 mg/m^2^ D1,2,3), cytarabine (100mg/m^2^/d CI D1-7), and gemtuzumab ozogamicin (6 mg/m^2^ D4) (“DA+GO”). Rates of cytomorphological complete remission (69% and 70%), 5-year relapse-free survival (43% and 42%), and 5-year overall survival (46% and 50%) have previously been reported as not different between DA and DA+GO arms respectively.^18^

Utilizing samples and clinical data from patients treated on the S0106 trial, we explored the utility of MRD to predict treatment outcomes by both MFC and NGS. Bone marrow specimens obtained prior to treatment and at time of CR underwent centralized, prospective assessment of MRD using a 3 tube, 10-color MFC assay.^19^ Banked samples from a total of 67 patients were available at diagnosis and CR after first induction, and 62 patients with trackable variants identified using a 29 gene NGS panel at diagnosis underwent retrospective genomic analysis with error-corrected duplex sequencing (DS) for MRD at time of CR.

## Methods

### Patients

Archival bone marrow (BM) aspirates or peripheral blood (PB) from 67 patients enrolled on the SWOG trial S0106 (NCT00085709) were available for this study. A total of 62 patients were selected for MRD analysis if they 1) achieved a first morphological complete remission (CR) with protocol induction therapy, 2) had both diagnosis and remission samples after first induction, 3) had central flow cytometry results on their remission marrow, and 4) had a variant detected at diagnosis for tracking in remission. Samples described in this manuscript were collected at time of first morphologic CR, but if CR samples were not available the first sample collected after achieving CR was used. BM (n=56) and PB (n=6) remission samples were collected a median of 34 days (range 25 to 162) post-randomization and a median of 0 days (range -6 to 121) from clinically defined remission. Institutional review boards of participating sites approved all protocols, and patients were treated according to the Declaration of Helsinki.

### DNA Extraction and Quantification

Genomic DNA (gDNA) from cryopreserved patient bone marrow or peripheral blood mononuclear cells was extracted with the Qiagen PureGene kit. gDNA from a separate young, healthy donor was extracted from a Leukopak purchased from AllCells using an Agilent Genomic DNA extraction kit. For technical spike-in mixtures, gDNA from HCC827, AN3-CA, SW1271, MDA-MB-453, SW48, HCT-15, and SW620 cells was purchased from ATCC and gDNA from OCI-AML3, MOLM- 14-L1, and K-562 cells was a gift from Dr. Jerald Radich. All gDNA concentrations were quantified with the Qubit dsDNA High Sensitivity kit and quality assessed on an Agilent TapeStation 2200 using a Genomic DNA Screen Tape.

### Duplex Sequencing

Retrospective targeted DNA sequencing of 29 genes recurrently mutated in adult AML was performed on paired diagnostic and remission bone marrow or peripheral blood samples utilizing the TwinStrand Duplex Sequencing™ AML-29 Panel (**Supplementary Table 1**). Non-error corrected sequencing was performed on diagnostic samples (500ng gDNA) and error-corrected duplex sequencing (DS) was performed on remission samples (1μg gDNA). DS was performed essentially as described.^20^ Briefly, gDNA was sheared to a peak fragment size of 300 bp using a Covaris ultrasonicator. End repair, A-tailing and DuplexSeq™ adapter ligation were performed prior to library conditioning with a cocktail of glycosylases to remove damaged DNA molecules prior to amplification. Following indexing PCR, libraries were hybridized with biotinylated 120-mer DNA probes and purified with streptavidin magnetic beads. Following washes additional PCR was performed, followed by another round of hybridization, capture, washes, and final PCR. Libraries were sequenced using paired-end 150bp sequencing on an Illumina NextSeq 500 (diagnostic samples) or a NovaSeq 6000 (remission samples). FASTQ files are available in the NCBI Small Reads Archive (SRA) (Accession: PRJNA945188).

For technical assessment of the 29 gene panel with the DS assay, mutant cell line DNA was spiked into healthy donor DNA at predicted VAFs ranging from 1.0×10^−2^ to 3.9×10^−6^. Mutant DNA samples harboring a total of 21 unique variants across 13 genes were combined into mutation mixes. Fifteen single nucleotide variants (SNV) were combined into an “SNV mix” and 4 insertions/deletions (indel) were combined into an “indel mix”. A serial dilution of *FLT3*-ITD (21 bp) and *NPM1* insertion (4 bp) variants was also generated (1%, 0.1%, 0.01% and 0.003% VAF for each). Four replicate libraries were prepared for each mutation mix, with 1.5µg gDNA input for each library except 50ng for 1% *FLT3/NPM1* and 250ng for 0.1% *FLT3/NPM1*, and pure healthy donor DNA at each DNA input mass. Expected VAFs were based on COSMIC reported zygosities and dilution factors and were adjusted based on DS analysis of SNVs in pure cell line DNA and 1:1 mixes of cell line and healthy donor DNA. Libraries were prepared as above and sequenced on a NovaSeq 6000.

### Bioinformatics

Alignment, duplex consensus sequence generation, and variant calling were performed as described.^20^ For each patient, potential germline variants were identified and excluded from the analysis if the VAF was ≥ 35% at both diagnosis and remission, or ≥ 40% at either time point and a gnomAD allele frequency ≥ 0.05. Somatic variants present at diagnosis were classified as potentially deleterious if computationally predicted as such and with a VAF ≥ 5% (≥ 1% for *FLT3*- ITD/*NPM1* insertions). Somatic variants in remission followed the same classification rules for deleterious impact and required an alternative depth of ≥ 2 (≥ 1 for *FLT3*-ITD/*NPM1* insertions detected at diagnosis). All remaining variants were manually curated for pathogenicity. MRD by NGS was defined using conditions previously identified as prognostic.^7, 11^

### Multiparametric Flow Cytometry

Bone marrow samples collected at diagnosis and remission were analyzed for MRD using a 3 tube, 10-color multiparameter flow cytometry assay with a sensitivity of 0.1% in most cases; data and details of which were reported previously.^19^

### Statistics

Morphologic complete remission was defined per contemporary consensus criteria definitions and required count recovery with absolute neutrophil count > 1,000 and platelets > 100,000. Time-to-event outcomes analyzed were overall survival (OS; event=death), relapse-free survival (RFS, event=relapse or death) and time to relapse (TTR; event=relapse, death in remission a competing event). All outcomes were measured from date of morphologic remission to date of event, with patients without event censored at date of last contact. Associations between residual disease and outcomes were assessed using Cox regression models (cause-specific model for TTR); model discrimination was assessed using C-statistics.

## Results

### Patient characteristics

The median age of the 62 patients in this study was 48 (range 18-60) (**Table 1**). Thirty-two patients were randomized to DA and 30 patients to DA+GO. At 5 years, the rate of non-relapse mortality (NRM) was 10%, relapse was 33%, relapse-free survival was 57%, and overall survival was 64% for the entire cohort (**Figure 1**). Overall patient demographics and clinical outcomes of the 62 patients analyzed in this study align with the full S0106 clinical trial cohort (**Supplementary Table 2**).

**Table 1:** Patient clinical characteristics.

| **Covariate** | **Patient cohort** |
| --- | --- |
| **Number of patients** | 62 |
| **Randomized arm** |  |
| DA | 32 (52%) |
| DA+GO | 30 (48%) |
| **Age** |  |
| Median (range) | 48 (18-60) |
| **Sex** |  |
| Female | 28 (45%) |
| Male | 34 (55%) |
| **Performance status** |  |
| 0-1 | 58 (84%) |
| 2-3 | 11 (16%) |
| **Cytogenetic risk** |  |
| Favorable | 13 (23%) |
| Intermediate | 30 (54%) |
| Adverse | 13 (23%) |
| Missing | 6 |
| **WBC ($\times 10^3$)** |  |
| Median (range) | 18.0 (0.2-214) |
| **Platelets ($\times 10^3$)** |  |
| Median (range) | 48.5 (10, 449) |
| **Hemoglobin (g%)** |  |
| Median (range) | 9.4 (3.5-13.6) |
| **Race** |  |
| Asian | 1 (2%) |
| Black | 4 (6%) |
| Native American/Alaskan | 1 (2%) |
| Pacific Islander | 0 |
| White | 54 (87%) |
| Unknown | 2 (3%) |
| **Specimen for sequencing** |  |
| Bone Marrow | 56 (90%) |
| Peripheral Blood | 6 (10%) |

WBC, white blood cell count; DA, daunorubicin and cytarabine; GO, gemtuzumab ozogamicin

**Figure 1.**
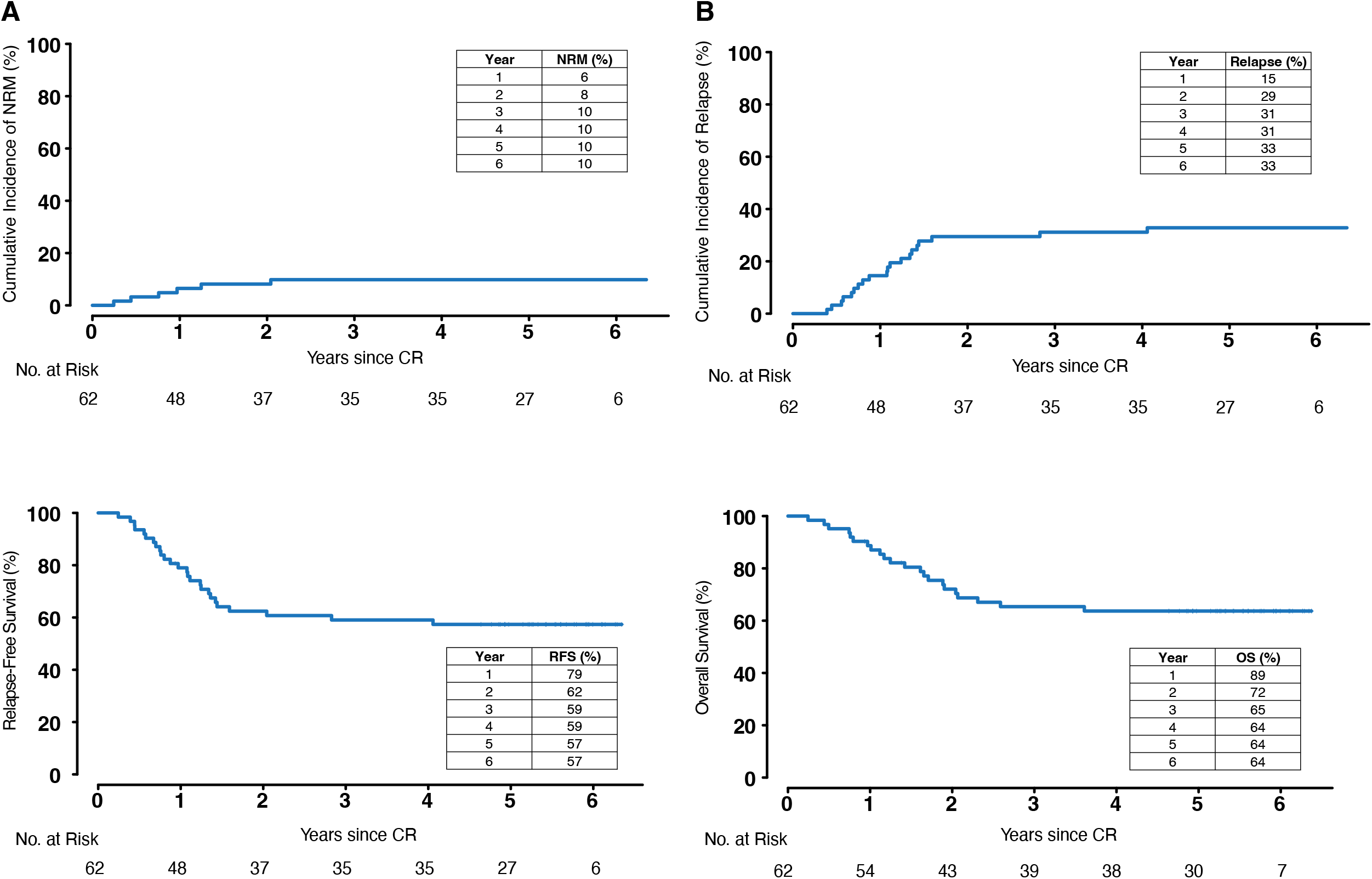
Clinical outcomes of S0106 AML patients analyzed for MRD. Rates of (A) non-relapse related mortality (NRM), (B) relapse, (C) relapse-free survival (RFS), and (D) overall survival (OS) are shown for the 62-patient cohort from the S0106 clinical trial analyzed for measurable residual disease (MRD) by duplex sequencing and multiparametric flow cytometry.

Targeted sequencing analysis of diagnostic samples at an average raw sequencing read depth of 279x utilizing a 29 gene panel identified a total of 172 potentially deleterious variants across the 62 patients. Variants had a median variant allele fraction (VAF) of 34% (range 1.4-91.5%) and were detected in 23 genes, with *FLT3* being the most frequently mutated (**Supplementary Table 3 and Supplementary Figure 1**). Patients had a median of 2 variants detected at diagnosis (range 1-9) that could be tracked in remission.

### Technical performance of DS

Technical performance of the 29 gene DS assay was assessed on contrived mutation mixes versus healthy donor DNA. An SNV mix containing 15 variants, an indel mix containing 4 variants, and 4 separate serial dilutions of a *FLT3-*ITD*/NPM1* mutant mix were analyzed, with predicted VAFs ranging from 1.0×10^−2^ to 3.9×10^−6^. Data combined from 4 replicate libraries per mix generated 135,065-142,707x mean duplex consensus molecular depth (from the 1.5 µg DNA input libraries), with max depths 186,645-196,896x. All expected variants were detected in the mutation mixes and the observed VAFs were significantly correlated with the predicted VAFs (r^2^ > 0.99, **Supplementary Figure 2**). When the 21 spike-in mutation positions were assessed in the pure healthy donor DNA, a total of 4 mutant allele counts were detected out of a total duplex molecular depth of 2,993,429x at the 21 spike-in sites, for a combined mutation frequency of 1.3×10^−6^. The highest single background VAF at a spike-in site in the pure healthy donor DNA was 1.3×10^−5^.

### Detection of residual variants in remission

DS of remission samples utilizing the same 29 gene panel at a median error-corrected duplex molecular depth of 27,996x (range 11,958x-35,131x) identified 82 diagnostic variants remaining in remission, with a median VAF of 0.059% (range 0.005-41.8%) (**Supplementary Table 3**).

Variants were detected in 18 genes, with *DNMT3A* being the most frequently mutated, followed by *NPM1* and *FLT3* (**Supplementary Figure 3**). Forty-three patients (69%) had at least one diagnostic variant detectable in remission, with a median of 2 residual variants per positive patient (range 1-5). Residual diagnostic variants in remission had a median 2.60 (range 0.06-3.96) log^10^ reduction in VAF. Not surprisingly, mutations in *DNMT3A* and *TET2*, genes commonly associated with clonal hematopoiesis, showed the least change in VAF between diagnosis and remission (median 1.23 (range 0.06-3.31) and 1.32 (range 1.23-2.29) log^10^ reduction, respectively). Mutations in *FLT3* showed the greatest change in VAF, median 3.12 (range 1.5-3.8) log^10^ reduction.

### MRD as defined by flow cytometry

MFC analysis of bone marrow collected at the time of remission using a 3 tube, 10-color assay identified MRD in 10 (16%) patients (**Figure 2**) and the median MRD level was 0.25% (range 0.002-6.2%) (**Supplementary Table 4**). Patients who were MFC MRD positive had increased rates of relapse (50% vs 30% at year 5; HR, 2.4; 95% CI, 0.9-6.7; P=0.087) and decreased rates of relapse-free survival (40% vs 61% at year 5; HR, 2.2; 95% CI, 0.9-5.4; P=0.095) and overall survival (40% vs 68% at year 5; HR, 2.5; 95% CI, 1.0-6.3; P=0.059) compared to patients that were MFC MRD negative (**Figure 3A, Table 2**). While not statistically significant in this subset of S0106 patients, these results, including the magnitude of the HRs, are in line with the significant findings previously published for the larger cohort of S0106 patients analyzed by MFC.^19^

**Figure 2.**
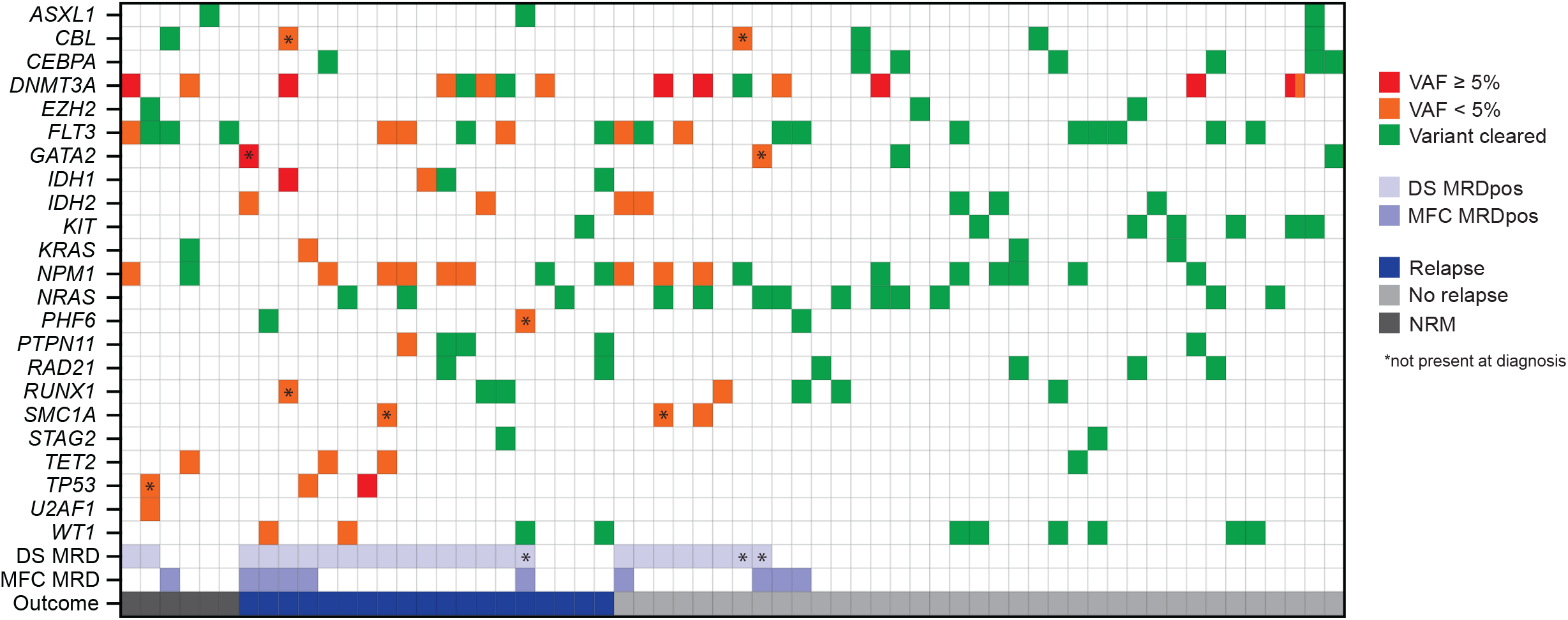
Mutational spectrum, MRD status, and clinical outcomes of patients in complete remission. The heatmap displays variants detected at diagnosis and the presence (divided into variant allele fraction (VAF) ≥ or < 5%) or absence at the time of complete remission by duplex sequencing (DS), DS measurable residual disease (MRD) status, multiparametric flow cytometry (MFC) MRD status, and clinical outcome at 5 years (relapse, no relapse, or non-relapse mortality (NRM). The presence of a mutation within a gene is denoted in the heatmap, with the color corresponding to the highest VAF within each gene per patient. Variants identified in remission that were not identified at diagnosis are also marked (*).

**Figure 3.**
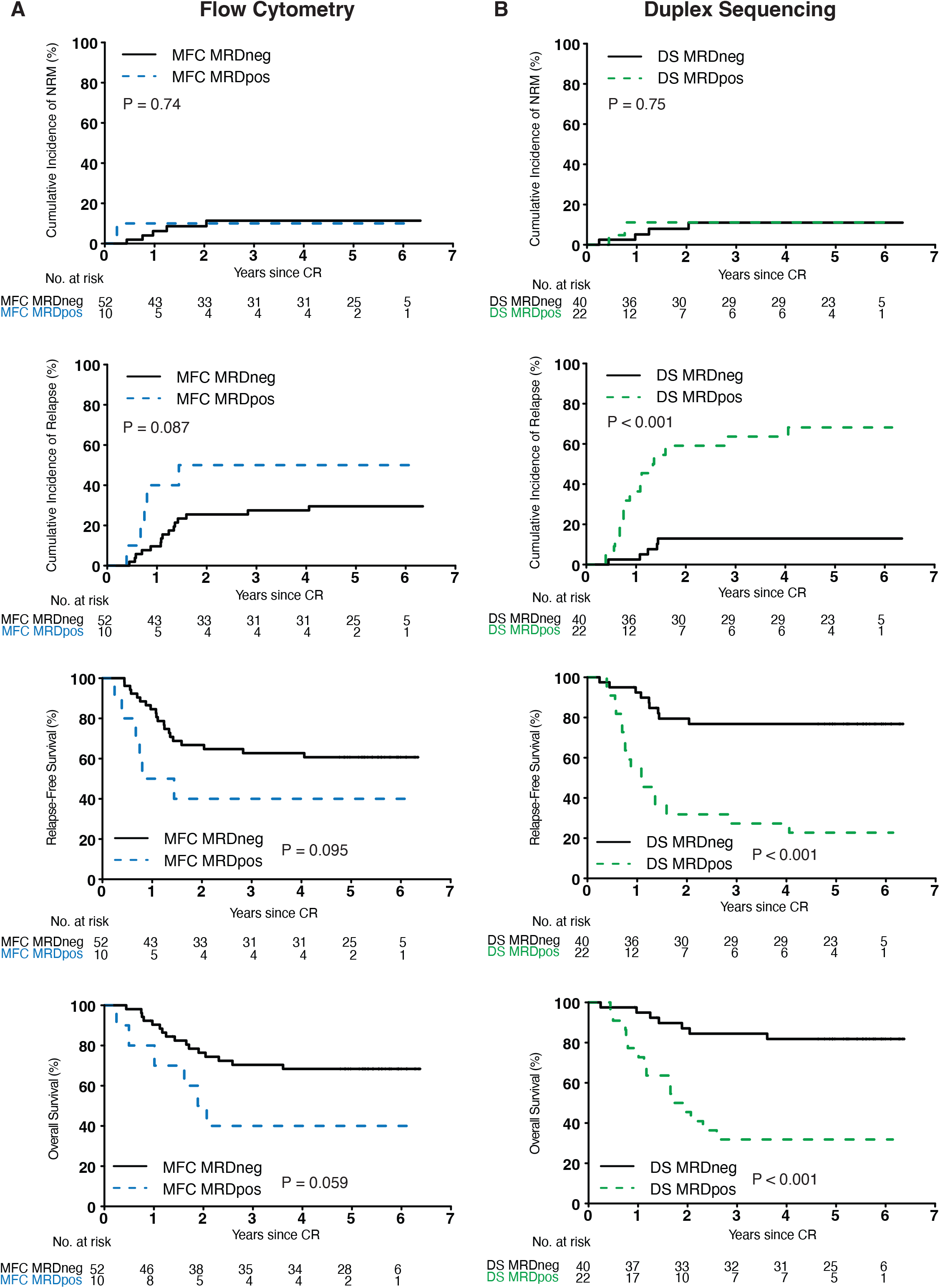
Impact of MRD status on clinical outcomes. Rates of non-relapse mortality (NRM), relapse, relapse-free survival, and overall survival are shown based on measurable residual disease (MRD) status as determine by (A) multiparametric flow cytometry (MFC) and (B) duplex sequencing (DS). Positive, pos; Negative, neg.

**Table 2.**
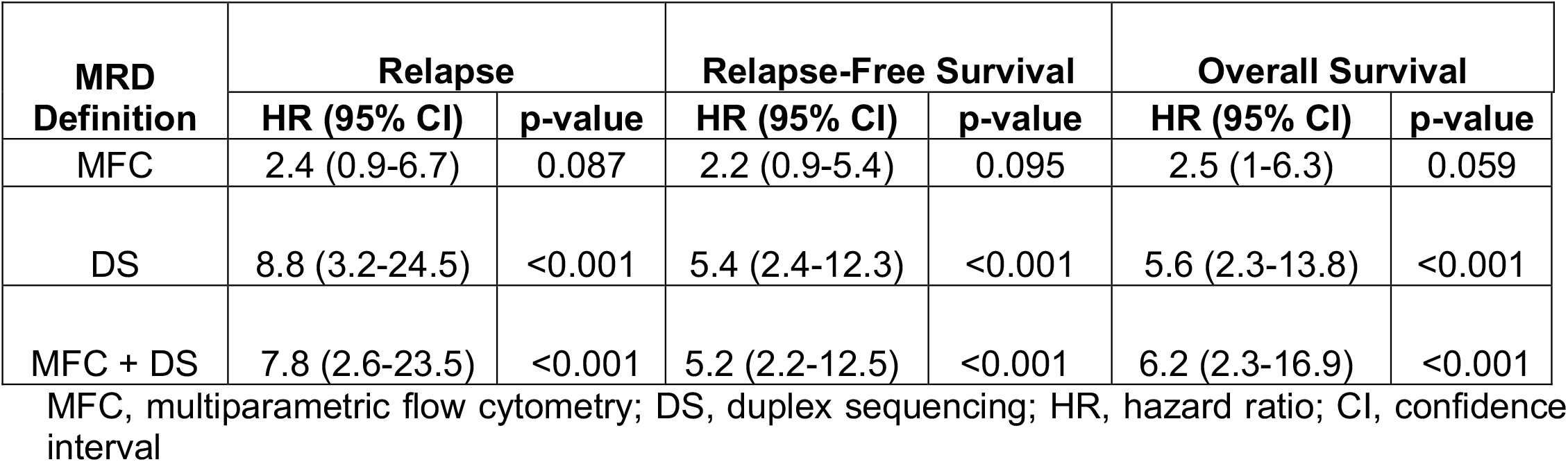
Univariate cox regression model for associations between MRD definitions and clinical outcomes.

### MRD as defined by detection of residual diagnostic variants by DS

We defined DS test positivity utilizing criteria previously demonstrated to be prognostic for AML MRD by NGS^7, 11^, which included non-*DTA* (*DNMT3A, TET2, ASXL1*) time-of-diagnosis mutations with a VAF ≥ 0.1% and/or an *FLT3*-ITD/*NPM1* VAF ≥ 0.01% (**Figure 2**). Using this definition, 22 patients (35%) were DS MRD positive. Compared to MFC, DS MRD provided a superior prediction of clinical outcomes, such that patients who were DS MRD positive had significantly increased rates of relapse (68% vs 13% at year 5; HR, 8.8; 95% CI, 3.2-24.5; P<0.001) and decreased rates of relapse-free survival (23% vs 77% at year 5; HR, 5.4; 95% CI, 2.4-12.3; P<0.001) and overall survival (32% vs 82% at year 5; HR, 5.6; 95% CI, 2.3-13.8; P<0.001) compared to patients that were DS MRD negative (**Figure 3B, Table 2**).

Additional criteria for defining MRD by DS were also explored, including investigating the presence of any residual diagnostic variant and filtering based on VAF, gene, and VAF log^10^ reduction relative to diagnosis (**Supplementary Figure 4 and Supplementary Table 5**). While a naïve definition of AML MRD as the detection of any residual diagnostic variant in remission was not associated with statistically significant difference in rates of relapse or survival (**Supplementary Figure 4 and Supplementary Table 5**), the addition of VAF cutoffs, removal of mutations in genes associated with clonal hematopoiesis (*DTA*), and limiting calls to variants with no more than a log^10^ reduction of 2 between diagnosis and remission all resulted in statistically significant increased rates of relapse and decreased overall survival and relapse-free survival compared to patients testing negative, but none outperformed the criteria previously established as prognostic.

### MRD as defined by *de novo* detection of deleterious variants by DS

We also explored the value of detecting AML-associated variants in remission that were not detected at the time of diagnosis. Utilizing the same variant filtering as defined above but agnostic to variant status at diagnosis, we identified 12 additional variants across 9 patients with a median VAF of 0.24% (range 0.08-15.1%) (**Supplementary Table 4**). This resulted in 3 additional patients being defined as DS MRD positive, for a total of 25 (40%) patients (**Figure 2**). Use of this NGS MRD definition agnostic to diagnostic variants provided a similar prediction of clinical outcomes to that of the initial prognostic criteria, such that patients who were DS MRD positive had significantly increased rates of relapse (64% vs 11% at year 5; HR, 8.7; 95% CI, 2.9-26.1; P<0.001) and decreased rates of relapse-free survival (28% vs 78% at year 5; HR, 4.8; 95% CI, 2.1-11.1; P<0.001) and overall survival (36% vs 83% at year 5; HR, 5.4; 95% CI, 2.1-13.8; P<0.001) compared to patients that were DS MRD negative (**Supplementary Figure 4F, Supplementary Table 5**).

### Comparison of MRD detection by MFC versus DS

Next, we examined the differences between MFC and DS MRD calls. Of the 62 patients analyzed, 5 (8%) were called positive and 35 (56%) were called negative for MRD by both MFC and DS (**Figure 4A**). Five of the 10 (50%) patients called MRD positive by MFC were called negative by DS and 17 of the 22 (77%) patients called MRD positive by DS were called negative by MFC.

**Figure 4.**
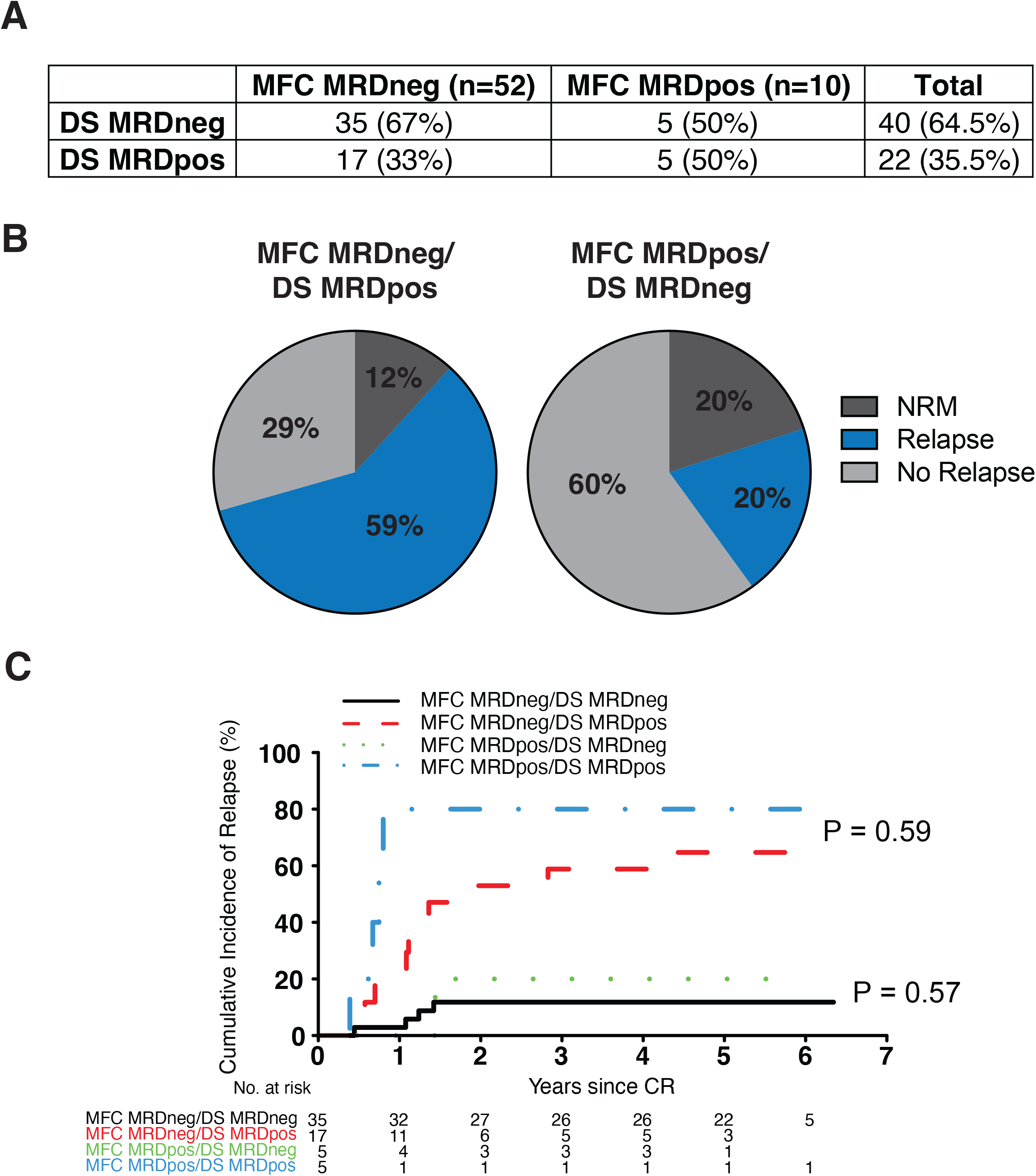
Analysis of discordant MRD results by duplex sequencing and flow cytometry. (A) Number and percentage of patients called MRD positive (pos) versus MRD negative (neg) by duplex sequencing (DS) versus multiparametric flow cytometry (MFC). (B) Clinical outcomes (non-relapse mortality (NRM), relapse, or no relapse) of MFC MRD versus DS MRD discordant cases. (C) Rates of relapse for patients grouped by MRD status as defined by MFC MRD and DS MRD. Complete remission, CR; Number, No.

Comparing clinical outcomes of the discordant cases revealed that 59% of patients called MFC MRD negative/DS MRD positive relapsed, while only 20% of patients called MFC MRD positive/DS MRD negative relapsed (**Figure 4B**). While patients defined as MRD positive by both MFC and DS had the highest rate of relapse (80% at year 5), there was no significant difference in rates of relapse between DS MRD positive/MFC positive and DS MRD positive/MFC negative (80% vs 65% at year 5, cause-specific P=0.59) or DS MRD negative/MFC positive and DS MRD negative/MFC negative (20% vs 12% at year 5, cause-specific P=0.57), indicating DS MRD was the main driver of outcomes prediction (**Figure 4C, Supplementary Figure 5A**).

Looking closer at the disease burden in the discordant cases that experienced relapse, we found that the median VAF of variants identified in the DS MRD positive/MFC positive patients was 25 times higher than those identified in the DS MRD positive/MFC negative patients (1% vs 0.04%). Additionally, 5 out of the 10 (50%) DS MRD positive/MFC negative patients that experienced relapse had a residual variant in *NPM1* detected, compared to none in the DS MRD positive/MFC positive patients.

Furthermore, the addition of MFC to the DS MRD definition did not significantly improve outcome predictions. While patients who were MFC and/or DS MRD positive had significantly increased rates of relapse (59% vs 12% at year 5; HR, 7.8; 95% CI, 2.6-23.5; P<0.001) and decreased rates of relapse-free survival (30% vs 79% at year 5; HR, 5.2; 95% CI, 2.2-12.5; P<0.001) and overall survival (37% vs 85% at year 5; HR, 6.2; 95% CI, 2.3-16.9; P<0.001) compared to patient who were MFC and DS MRD negative, this did not significantly differ from DS MRD alone (**Table 2, Supplementary Figure 5B**).

We assessed the ability of covariates to predict relapse in individual patients. Baseline clinical characteristics (including age, performance status, and cytogenetics) yielded a C-statistic of 0.66. Inclusion of MFC MRD status did not improve the discrimination of the model with a C-statistic of 0.67, while inclusion of DS MRD status did improve the model discrimination yielding a C-statistic of 0.77.

### Impact of DS MRD status and treatment regimen on clinical outcomes

Finally, we examined the impact of DS MRD status and patient randomization to DA versus DA+GO on clinical outcomes. In concordance with results from the full S0106 cohort^18^, the subset of 62 patients in this study showed no difference in rates of relapse between patients treated with DA versus DA+GO (35% vs 31% at year 5, P=0.62) (**Figure 5A**). Adding information on DS MRD status showed that patients who were DS MRD positive had significantly higher rates of relapse compared to patients that were DS MRD negative regardless of the treatment regimen (DA: 60% vs 12% at year 5, P=0.017, DA+GO: 86% vs 13% at year 5, P< 0.001) (**Figure 5B**). No difference was seen in rates of relapse between patients treated with DA versus DA+GO based on DS MRD status (DS MRD positive: 60% vs 86% at year 5, P=0.2, DS MRD negative, 12% vs 13%, P=0.98).

**Figure 5.**
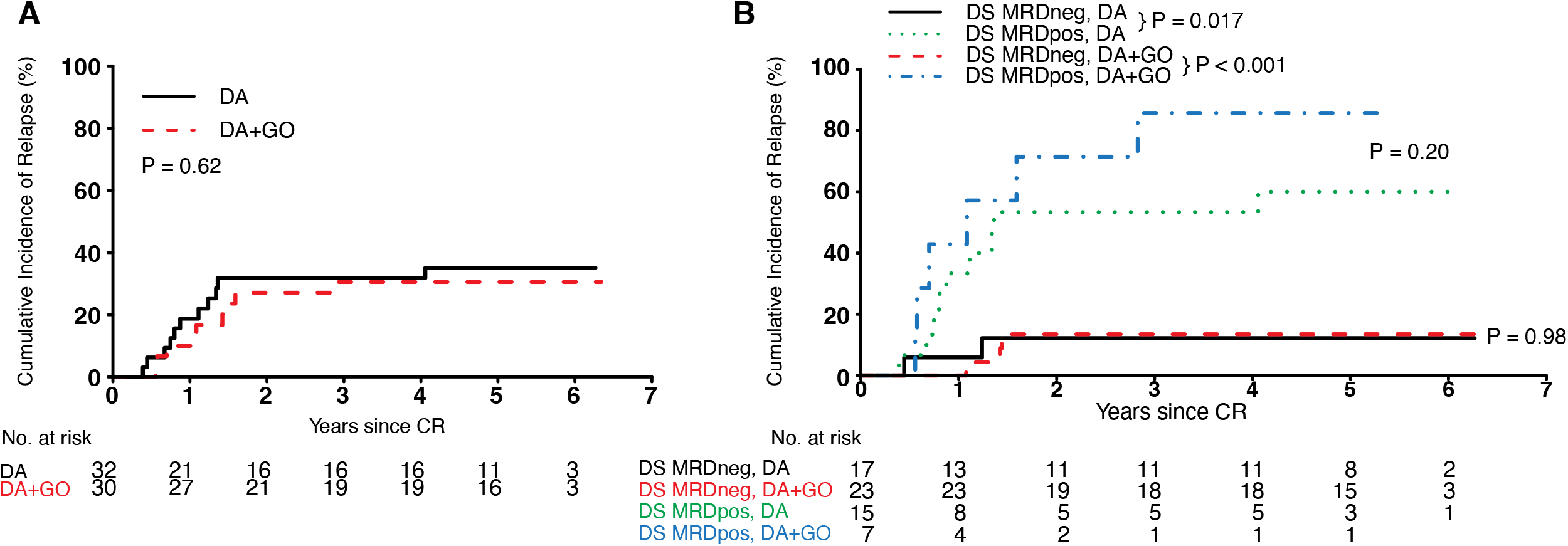
Impact of treatment randomization and DS MRD status on relapse. Rates of relapse for patients as defined by (A) treatment randomization to daunorubicin and cytarabine (DA) versus daunorubicin, cytarabine, and gemtuzumab ozogamicin (DA+GO) and (B) treatment randomization (DA or DA+GO) and duplex sequencing (DS) measurable residual disease (MRD) status. Positive, pos; Negative, neg.

## Discussion

MRD has been well established as a method for quantifying the antileukemic effect of interventional therapies, but implementation in the clinic has thus far been limited for AML. MFC has been widely used for AML MRD detection, but concerns exist over inter-lab variability which could hinder widespread applicability of this technique. NGS for AML MRD detection could be more amenable to decentralized clinical testing and has been shown to outperform decentralized flow cytometry in the context of *FLT3*-ITD and *NPM1* mutated AML.^11^ Utilizing a subset of 62 AML patients treated on the S0106 phase 3 randomized trial of DA versus DA+GO induction chemotherapy, we compared the performance of MRD detection by high quality, centralized MFC and ultra-sensitive DS across a broad 29 gene panel to predict clinical response in first remission and found the latter to broadly have superior outcome-predicting performance.

Application of NGS for AML MRD detection has varied across the literature, and questions remain regarding the impact of assay sensitivity, gene targets, variant status at diagnosis, and the applicability across patients with diverse baseline genetics.^21, 22^ In the 67 patients screened in this study, we found that 62 (93%) had at least one variant present at diagnosis in the gene panel examined that could be tracked by NGS. The mutations identified spanned 23 genes, representing a broad set of AML MRD targets. The highly sensitive DS assay detected residual mutations at some level in most patients, rendering the naïve designation of MRD positivity clinically uninformative. However, application of previously established, clinically relevant variant filtering conditions, including VAF thresholds well above the assay limit of detection and removal of less informative genes (*DNMT3A, TET2, ASXL1*) associated with clonal hematopoiesis^23, 24^, was highly predictive of adverse clinical outcomes. These results highlight the importance of establishing informed guidelines for interpreting the presence of molecular MRD in the clinical setting. Additionally, we found that utilizing these filtering criteria remains highly predictive when agnostic to diagnostic variants. Therefore, the DS assay may have utility even when a diagnostic sample is not available. Future studies are needed to assess clinically relevant VAF thresholds at later treatment time points where residual disease may be present at a lower level.

In comparison to MFC, DS was significantly better at stratifying patients at risk of adverse clinical outcomes. Additional prognostic value was not seen when combining MRD detection by DS and MFC. Of the DS MRD positive patients that relapsed, the median VAF of patients with MRD also detected by MFC was 25x higher than MFC negative patients and all relapses occurred within the first year, indicating that this subset of patients had a higher disease burden at the time of clinical remission. Of the DS MRD positive patients that relapsed but were MFC negative, 50% (n=5) had residual *NPM1* mutations, in contrast to none in the double positive group. *NPM1*-mutated AML characteristically has absent/low CD34 expression with heterogeneity seen in the observed leukemia-associated immunophenotypes^25, 26^, making it uniquely challenging to track by MFC. This combined with increased assay sensitivity could explain most of the discrepant results and improved prognostic power of DS.

The S0106 phase 3 clinical trial found that randomization of AML patients to DA versus DA+GO induction chemotherapy provided no significant difference in clinical outcomes. One potential value of MRD testing is to provide a surrogate endpoint to predict long-term patient response, allowing for faster drug development/approval and to identify patients in need of additional therapy versus those who do not. In this cohort we found that DS MRD was able to predict clinical relapse, with no significant difference for the prognostic implications of MRD status seen in patients who received DA versus DA+GO, indicating that DS MRD status at remission could serve as an appropriate surrogate biomarker. Follow-up studies are needed to confirm the applicability of this technology as a definitive endpoint in a clinical trial setting.

Limitations of this study include (1) the small sample size, (2) the retrospective nature of the DS MRD analysis, (3) the comparison to an early generation MFC assay, and (4) the age of the S0106 study potentially limiting comparability to contemporary AML standard of care. The findings of this study need to be confirmed in a larger cohort using prospective analysis by both DS and a more modern MFC MRD assay. Additionally, 5 of the 67 (7%) patients screened for this study were excluded due to lack of a mutation detected at diagnosis available for tracking by DS in the panel used. However, these patients did have cytogenetic abnormalities present. Whole exome or genome sequencing at diagnosis could inform individualized MRD panels, targeting a combination of recurrently mutated genes, novel variants, and structural alterations. Future works needs to be done exploring the use of patient personalized MRD targets to expand applicability to all patients.

In conclusion, we provide evidence that in a group of genetically diverse *de novo* adult AML patients randomized to DA versus DA+GO induction chemotherapy that ultra-sensitive detection of residual variants by DNA sequencing in the bone marrow or blood at the time of first complete remission can outperform centralized, high-quality multiparametric flow cytometry in identifying patients at high risk of adverse clinical outcomes and predicting patient clinical response to treatment.

## Data Availability

All raw sequencing data produced are available online at the NCBI Small Reads Archive (Accession: PRJNA945188)

## Acknowledgements

This work was supported in part by the Intramural Research Program of the National Heart, Lung, and Blood Institute (CSH); National Cancer Institute of the National Institutes of Health under Award Number R44CA233381 (JS); and National Cancer Institute CA175008, 180888, 180819, 233338, and 233381 (JPR). The content is solely the responsibility of the authors and does not necessarily represent the official views of the National Institutes of Health.

