## Supplementary Information for "Quantification of measurable residual disease using duplex sequencing in adults with acute myeloid leukemia"


| | |
|---|---|
| Supplementary Figure 1. Detection of mutations at AML diagnosis. | Page 2 |
| Supplementary Figure 2. Technical assessment of the duplex sequencing assay. | Page 3 |
| Supplementary Figure 3. Detection of residual diagnostic mutations in remission. | Page 4 |
| Supplementary Figure 4. Association of different DS MRD status on clinical outcomes. | Page 5 |
| Supplementary Figure 5. Association of DS and/or flow cytometry MRD status on clinical outcomes. | Page 8 |
| Supplementary Table 1. Duplex sequencing panel target regions. | Page 9 |
| Supplementary Table 2. Patient clinical characteristics. | Page 10 |
| Supplementary Table 3. Variants detected at diagnosis and remission. | Page 11 |
| Supplementary Table 4. Flow cytometry MRD. | Page 14 |
| Supplementary Table 5. Univariate cox regression analysis for clinical outcomes based on various duplex sequencing MRD definitions. | Page 15 |

**Supplementary Figure 1. Detection of mutations at AML diagnosis.**
The (A) total number of variants per gene, (B) number of variants per patient (with or without including of *DNMT3A*, *TET2* or *ASXL1* (DTA) genes), and (C) variant allele fraction (VAF) of variants detected in diagnostic samples from 67 acute myeloid leukemia (AML) patients screened for inclusion in this study.

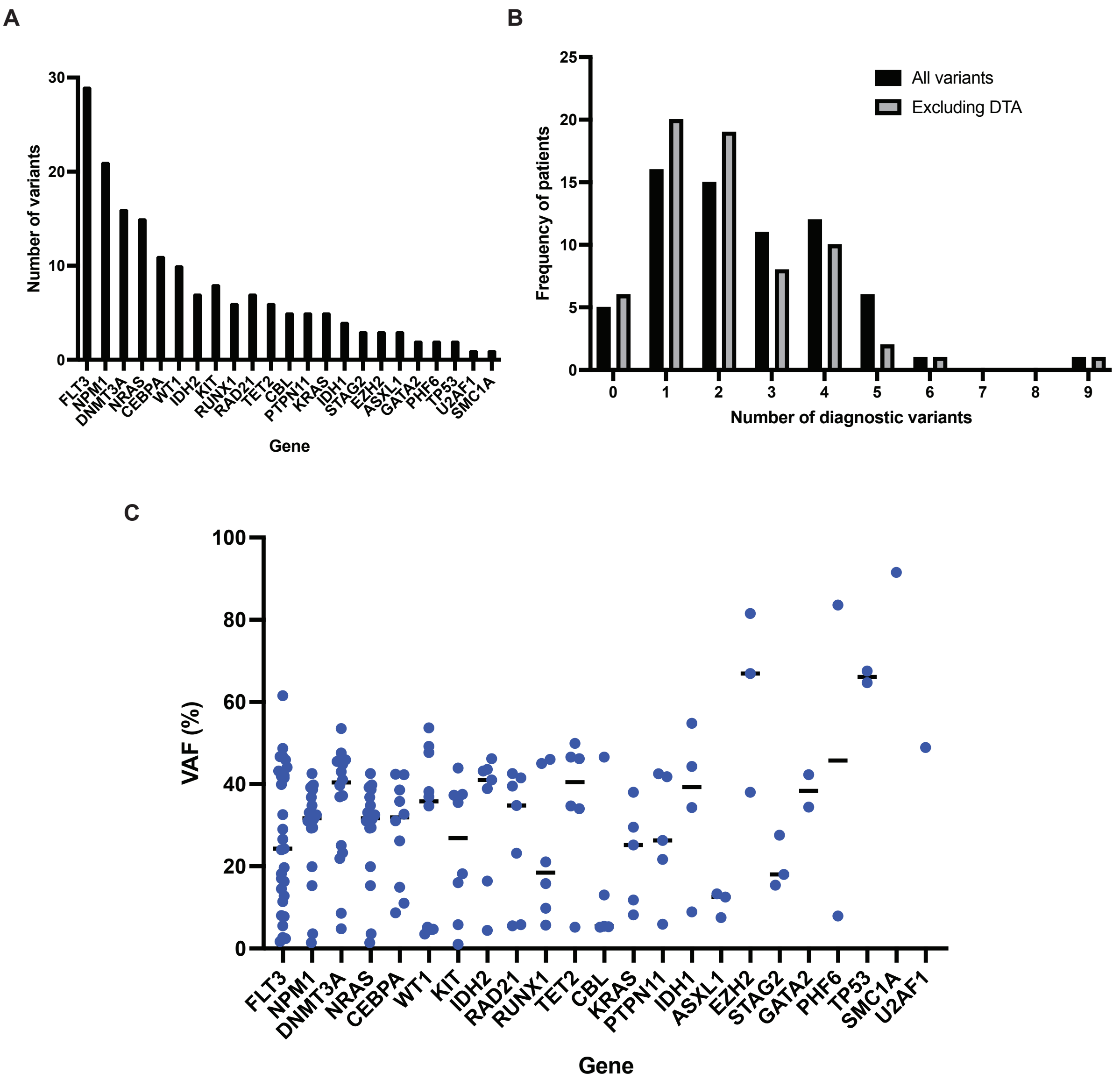

**Supplementary Figure 2. Technical assessment of the duplex sequencing assay.**
(A) Data from 4 technical replicate libraries were merged for each of the mutation mixes or negative controls. Mean and maximum panel-wide duplex molecular depth are plotted for each. (B) The expected vs. observed variant allele fractions (VAFs) are listed for each variant in the cell line DNA spike-in mixtures. *FLT3*-ITD and *NPM1* insTCTG each appear 4 times to reflect the serial dilutions of those mutations. (C) Observed vs. expected VAFs are plotted for each mutation mix, and correlations are calculated and inset. Error bars represent Wilson binomial 95% confidence intervals.

A

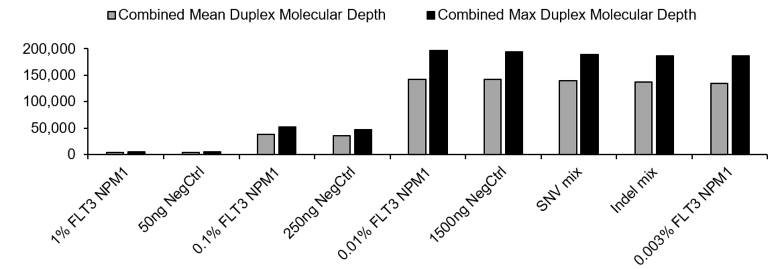


B

| Gene | Variant | Expected | Observed |
|---|---|---|---|
| *TP53* | C277F | 1.0x10$^{-2}$ | 9.4x10$^{-3}$ |
| *FLT3* | ITD | 1.0x10$^{-2}$ | 2.1x10$^{-2}$ |
| *NPM1* | insTCTG | 1.0x10$^{-2}$ | 4.1x10$^{-3}$ |
| *TP53* | Q136fs | 1.0x10$^{-2}$ | 5.5x10$^{-3}$ |
| *NRAS* | Q61R | 8.8x10$^{-3}$ | 7.1x10$^{-3}$ |
| *TET2* | R881G | 1.0x10$^{-3}$ | 7.4x10$^{-4}$ |
| *FLT3* | ITD | 1.0x10$^{-3}$ | 3.2x10$^{-3}$ |
| *NPM1* | insTCTG | 1.0x10$^{-3}$ | 8.4x10$^{-4}$ |
| *TP53* | V218fs | 1.0x10$^{-3}$ | 8.4x10$^{-4}$ |
| *TET2* | L1329M | 1.0x10$^{-4}$ | 1.0x10$^{-4}$ |
| *FLT3* | ITD | 1.0x10$^{-4}$ | 3.5x10$^{-4}$ |
| *NPM1* | insTCTG | 1.0x10$^{-4}$ | 9.4x10$^{-5}$ |
| *PTEN* | R130fs | 1.0x10$^{-4}$ | 6.6x10$^{-5}$ |
| *BRINP3* | C726* | 9.6x10$^{-5}$ | 5.8x10$^{-5}$ |

| Gene | Variant | Expected | Observed |
|---|---|---|---|
| *EZH2* | S734S | 5.7x10$^{-5}$ | 4.4x10$^{-5}$ |
| *TP53* | S90fs | 5.0x10$^{-5}$ | 3.2x10$^{-5}$ |
| *RAD21* | D116E | 5.0x10$^{-5}$ | 4.8x10$^{-5}$ |
| *HNRNPK* | Q182H | 4.9x10$^{-5}$ | 5.2x10$^{-5}$ |
| *DNMT3A* | M761I | 4.7x10$^{-5}$ | 2.1x10$^{-5}$ |
| *ASXL1* | W796C | 4.7x10$^{-5}$ | 1.8x10$^{-5}$ |
| *KRAS* | G13D | 4.6x10$^{-5}$ | 3.5x10$^{-5}$ |
| *BRINP3* | Q493K | 4.5x10$^{-5}$ | 4.3x10$^{-5}$ |
| *FLT3* | ITD | 3.0x10$^{-5}$ | 3.1x10$^{-5}$ |
| *NPM1* | insTCTG | 3.0x10$^{-5}$ | 3.5x10$^{-5}$ |
| *KRAS* | G12V | 3.0x10$^{-5}$ | 4.9x10$^{-5}$ |
| *TP53* | R273H | 2.4x10$^{-5}$ | 3.3x10$^{-5}$ |
| *RUNX1* | Q264R | 3.9x10$^{-6}$ | 9.1x10$^{-6}$ |

C

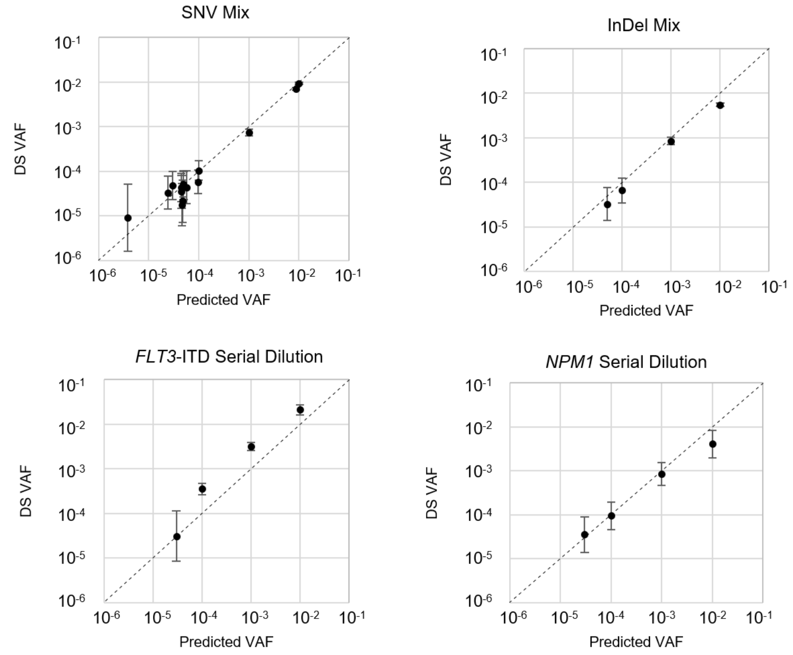

**Supplementary Figure 3. Detection of residual diagnostic mutations in remission.** The (A) total number of variants per gene, (B) number of variants per patient (with or without including of *DNMT3A*, *TET2* or *ASXL1* (DTA) genes), (C) variant allele fraction (VAF), and (D) -log[10] change in VAF between diagnosis and remission of diagnostic variants detected at the time of remission from 62 acute myeloid leukemia (AML) patients included in this study.

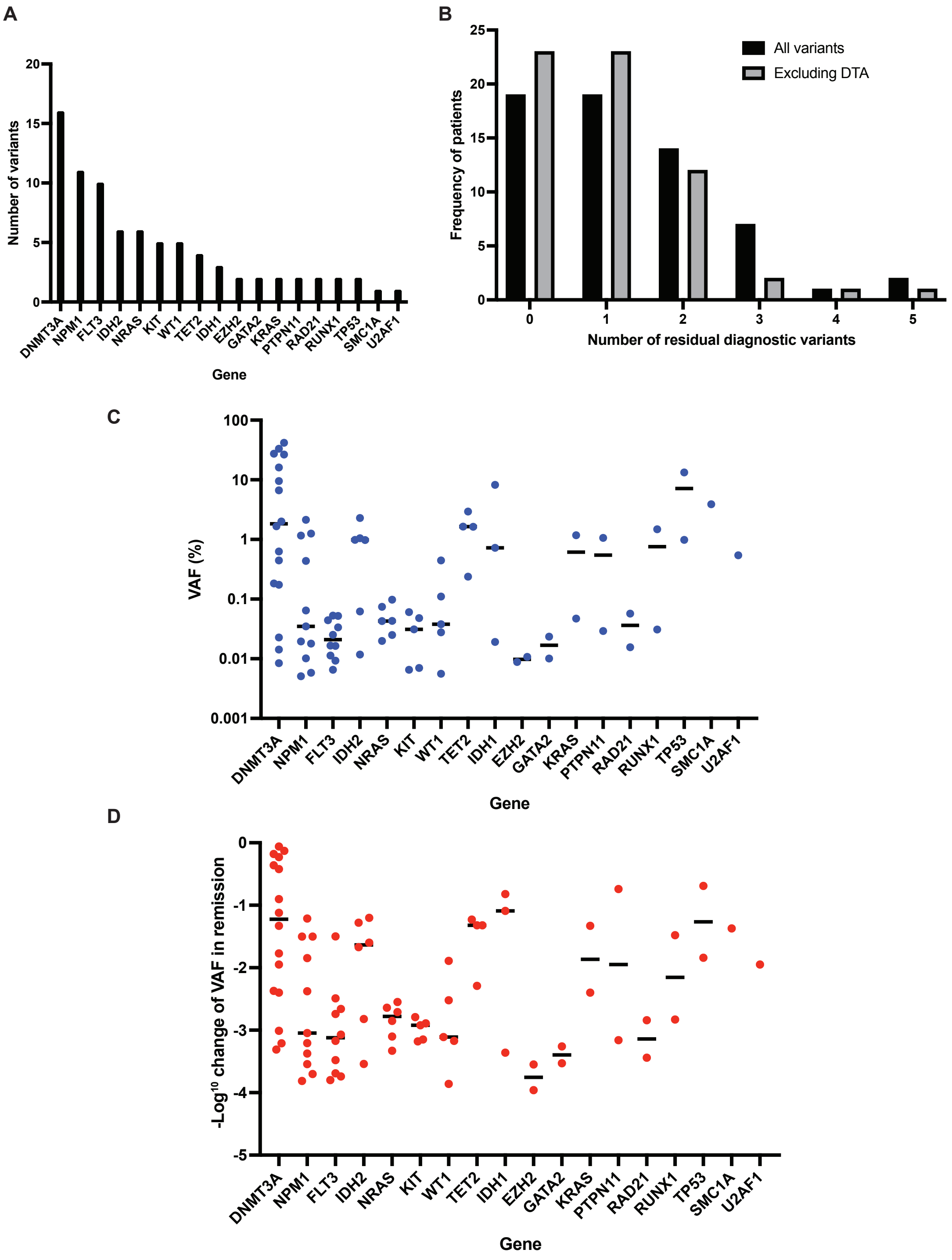

**Supplementary Figure 4. Association of different DS MRD status on clinical outcomes.** Rates of non-relapse mortality (NRM, top left), relapse (top right), relapse-free survival (bottom left), and overall survival (bottom right) are shown by remission DS MRD status as defined by: (A) any residual diagnostic variant (RDV), (B) RDV with VAF ≥ 0.1%, (C) RDV with VAF ≥ 0.1%, excluding *DNMT3A*, *TET2*, and *ASXL1* (DTA), (D) RDV with no greater than 2 log fold reduction in VAF , (E) RDV with no greater than 2 log$^{10}$ reduction in VAF between diagnosis and remission, excluding DTA, and (F) deleterious variant with VAF ≥ 0.1%, or ≥ 0.01% for *NPM1/FLT3*-ITD, excluding DTA, agnostic to diagnosis.

**Duplex Sequencing - Any Residual Diagnostic Variant**

A

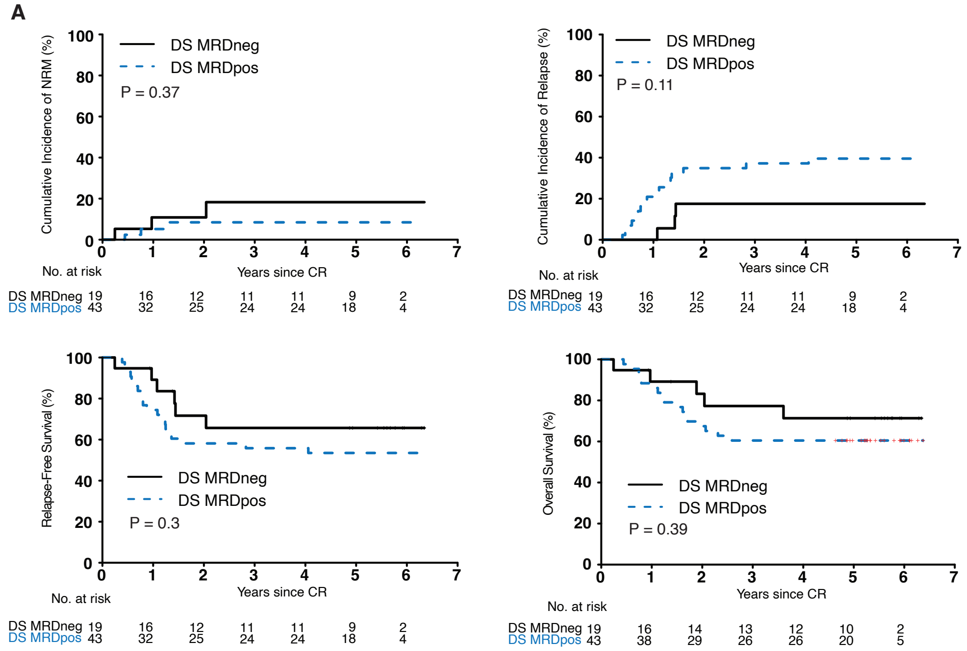


**Duplex Sequencing - Residual Diagnostic Variant with VAF ≥ 0.1%**

B

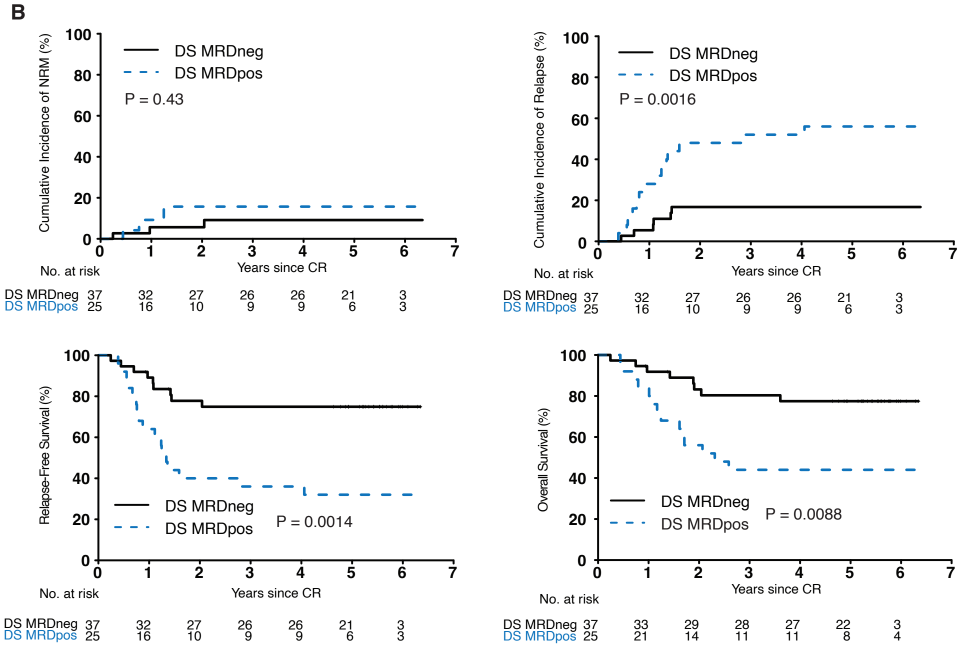

#### Duplex Sequencing - Residual Diagnostic Variant with VAF ≥ 0.1%, no DTA

C

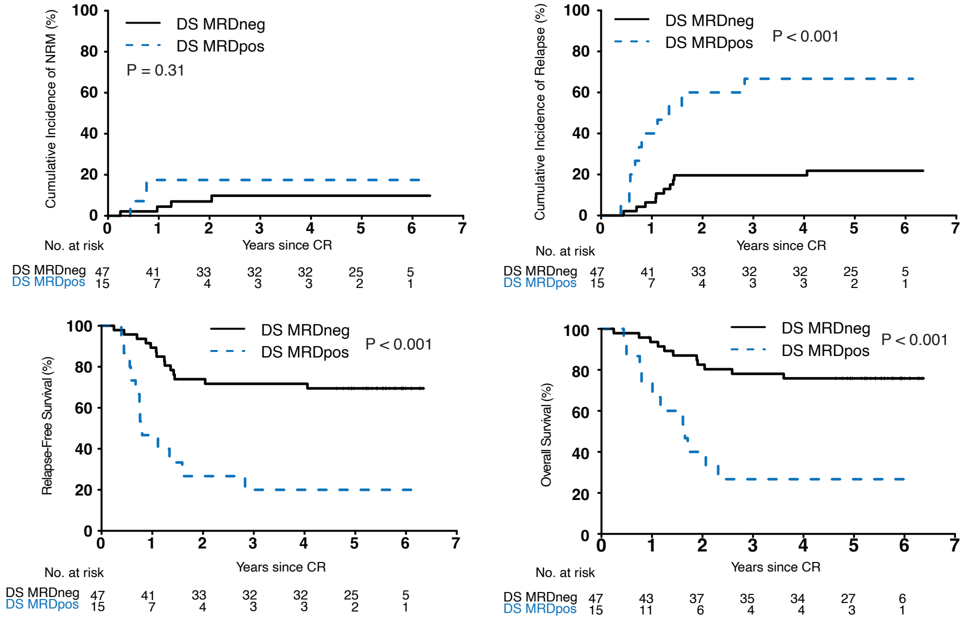


#### Duplex Sequencing - Residual Diagnostic Variant VAF ≤ 2 Log10 Decrease

D

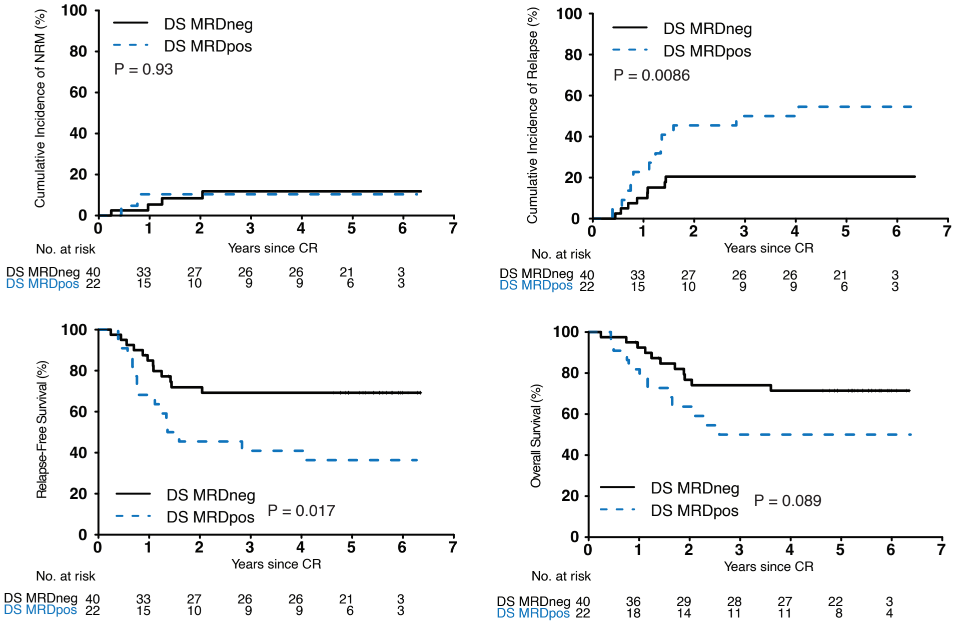

#### Duplex Sequencing - Residual Diagnostic Variant VAF ≤ 2 Log[10] Decrease, No DTA

E

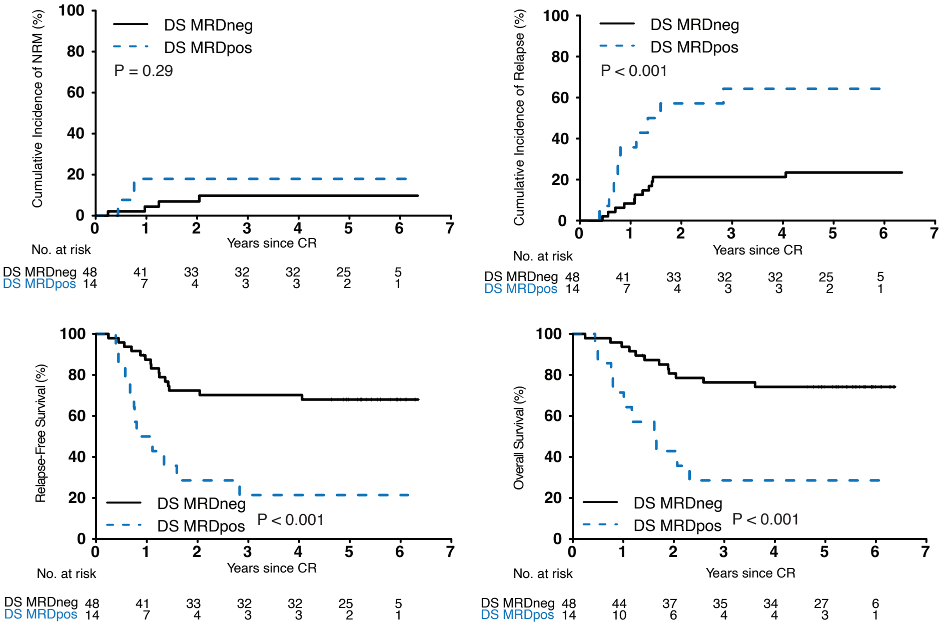


#### Duplex Sequencing - VAF ≥ 0.1% or ≥ 0.01% for *NPM1/FLT3*-ITD, No DTA, agnostic

F

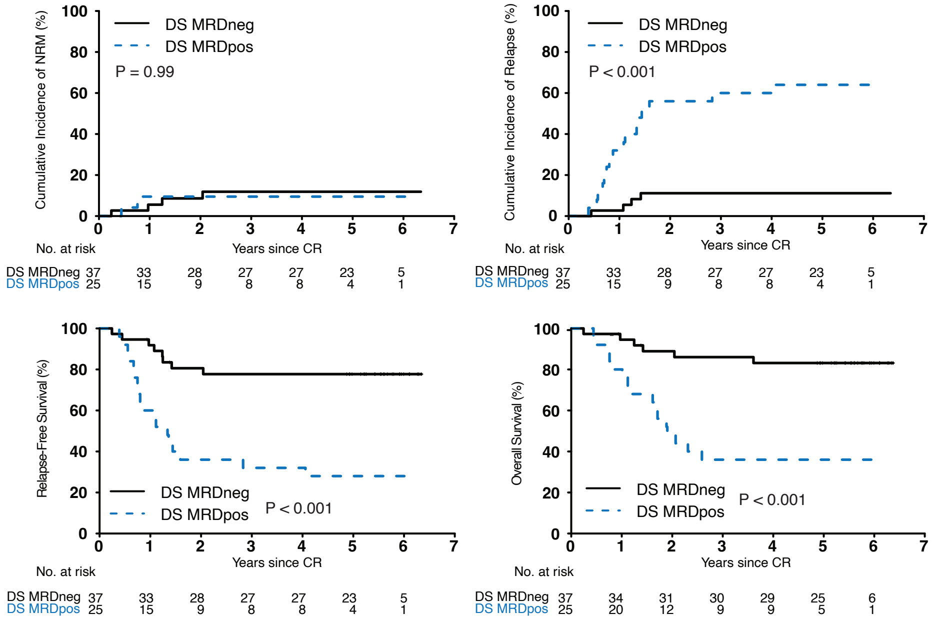

**Supplementary Figure 5. Association of DS and/or flow cytometry MRD status on clinical outcomes.**

Rates of non-relapse mortality (NRM, top left), relapse (top right), relapse-free survival (bottom left), and overall survival (bottom right) are shown by remission MRD status as determined by (A) DS and flow cytometry and (B) DS or flow cytometry.

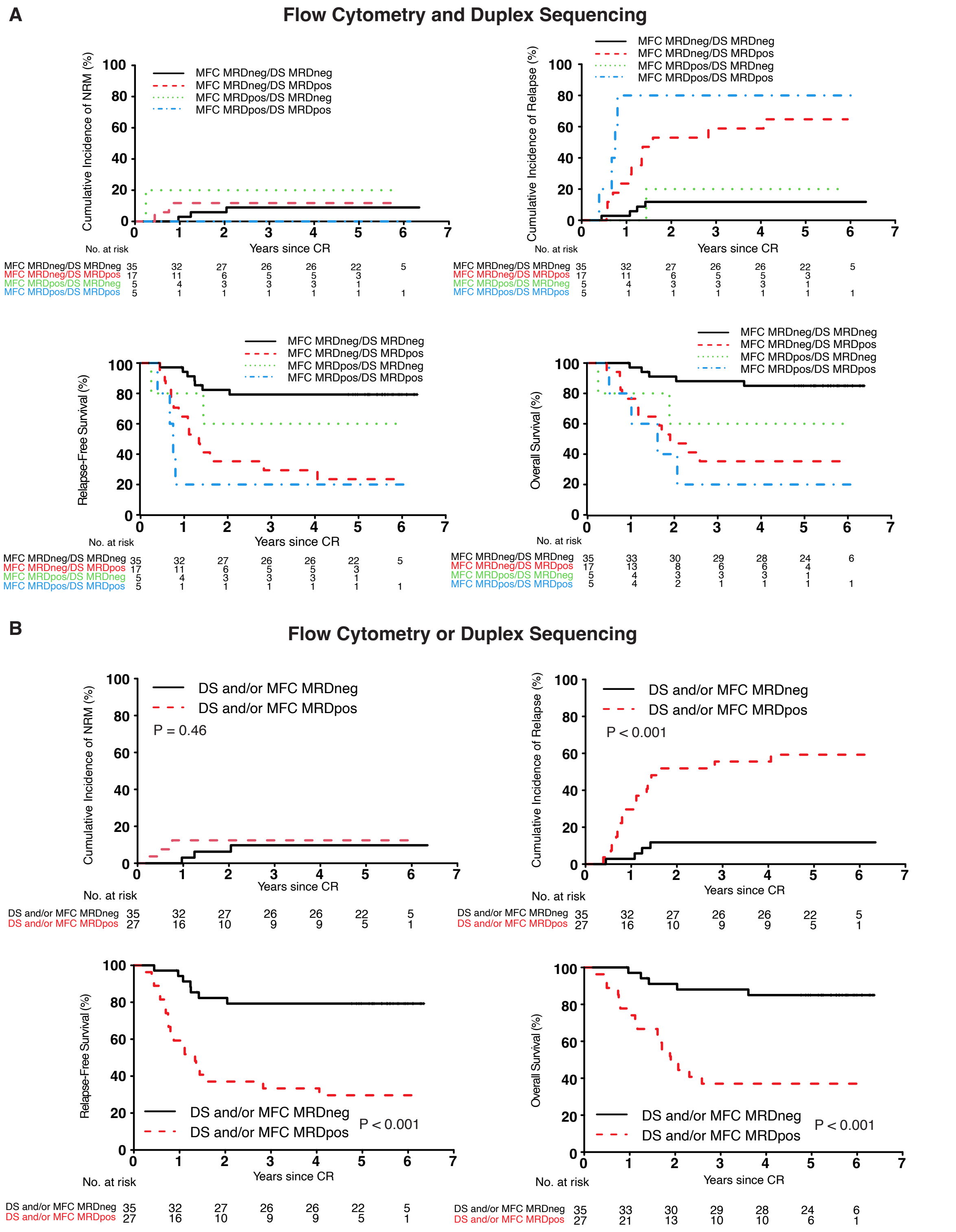

**Supplementary Table 1. Duplex sequencing panel target regions**

| Gene | Accession # | Target Region (amino acid) | Exon |
|---|---|---|---|
| *ASXL1* | NM_015338 | 363-1542 | 12-13 |
| *CBL* | NM_005188 | 366-477 | 8-9 |
| *CEBPA* | NM_001287424 | full CDS | 1 |
| *DNMT3A* | NM_022552 | 286-913 | 8-23 |
| *EZH2* | NM_004456 | 87-208, 244-302, 503-752 | 4-6, 8, 13-20 |
| *FAM5C (BRINP3)* | NM_199051 | 80-142, 396-767 | 3, 8 |
| *FLT3* | NM_004119 | 569-647, 807-847 | 14-15, 20 |
| *GATA2* | NM_032638 | 77-481 | 3-6 |
| *HNRNPK* | NM_002140 | 21-85, 173-215, 319-336, 371-453 | 4-6, 10, 12, 15-16 |
| *IDH1* | NM_005896 | 106-138 | 4 (partial) |
| *IDH2* | NM_002168 | 126-178 | 4 |
| *KIT* | NM_000222 | 412-448, 788-828 | 8, 17 |
| *KRAS* | NM_004985 | 1-96 | 2-3 |
| *MLL-X (KMT2A-X)* | NM_005933 (*KMT2A*) | *MLL* intron 9 | intron 9 |
| *MYH11-CBFB* | NM_022844 (*MYH11*) | *MYH11* intron 30, exon 31 | intron 30, exon 31 |
| *NPM1* | NM_002520 | 258-282, 283-295 | 10-11 |
| *NRAS* | NM_002524 | 1-96 | 2-3 |
| *PHF6* | NM_032458 | full CDS | 2-10 |
| *PTEN* | NM_001304717 | 258-337, 385-440 | 6, 8 |
| *PTPN11* | NM_002834 | 47-110, 484-533 | 3, 13 |
| *RAD21* | NM_006265 | full CDS | 2-14 |
| *RUNX1* | NM_001754 | full CDS | 2-9 |
| *SMC1A* | NM_006306 | 38-99, 447-515, 578-637, 687-732, 772-902, 1096-1145 | 2, 9, 11, 13, 15, 16-17, 22 |
| *SMC3* | NM_005445 | 184-268, 365-435, 656-705, 882-1035, 1100-1158 | 9-10, 13, 19, 24-25, 27 |
| *STAG2* | NM_001282418 | 42-128, 155-297, 436-472, 513-546, 578-675, 787-844, 892-1155 | 5-6, 8-10, 15, 17, 19-20, 25, 27-31 |
| *TET2* | NM_001127208 | full CDS | 3-11 |
| *TP53* | NM_000546 | full CDS | 2-11 + alt. exons |
| *U2AF1* | NM_006758 | 16-44 | 2 |
| *WT1* | NM_024426 | 372-523 | 7-10 |

CDS, coding sequence

**Supplementary Table 2. Patient clinical characteristics.**

| Covariate | Trial population | Achieved CR | MRD flow data available | Screened by NGS for molecular targets at diagnosis | Molecular target identified at diagnosis and DS data available |
|---|---|---|---|---|---|
| **Number of patients** | 595 | 416 | 184 | 67 | 62 |
| **Randomized arm** | | | | | |
| DA | 300 (50%) | 210 (50%) | 89 (48%) | 35(52%) | 32 (52%) |
| DA+GO | 295 (50%) | 206 (50%) | 95 (52%) | 32 (48%) | 30 (48%) |
| **Age** | | | | | |
| Median (range) | 47 (18-60) | 47 (18-60) | 47 (18-60) | 47 (18-60) | 48 (18-60) |
| **Sex** | | | | | |
| Female | 282 (47%) | 193 (46%) | 95 (52%) | 30 (45%) | 28 (45%) |
| Male | 313 (53%) | 224 (54%) | 89 (48%) | 37 (55%) | 34 (55%) |
| **Performance status** | | | | | |
| 0-1 | 518 (88%) | 367 (89%) | 155 (85%) | 56 (84%) | 58 (84%) |
| 2-3 | 74 (12%) | 47 (11%) | 28 (15%) | 11 (16%) | 11 (16%) |
| **Cytogenetic risk** | | | | | |
| Favorable | 77 (17%) | 67 (21%) | 34 (21%) | 16 (26%) | 13 (23%) |
| Intermediate | 263 (58%) | 196 (63%) | 92 (58%) | 31 (51%) | 30 (54%) |
| Adverse | 117 (26%) | 50 (19%) | 34 (21%) | 14 (23%) | 13 (23%) |
| Missing | 138 | 93 | 24 | 6 | 6 |
| **WBC ($x10^3$)** | | | | | |
| Median (range) | 11.4 (0.2-545) | 11.3 (0.2-545) | 13.5 (0.2-214) | 18.7 (0.2-214) | 18.0 (0.2-214) |
| **Platelets ($x10^3$)** | | | | | |
| Median (range) | 54 (2, 9300) | 54 (5, 9300) | 55 (5, 9300) | 44 (10-449) | 48.5 (10, 449) |
| **Hemoglobin (g%)** | | | | | |
| Median (range) | 9.1 (3.5-29.1) | 9.1 (3.5-28.5) | 9.2 (3.5-28.5) | 9.4 (3.5-13.6) | 9.4 (3.5-13.6) |
| **Race** | | | | | |
| Asian | 26 (4%) | 12 (3%) | 3 (2%) | 1 (2%) | 1 (2%) |
| Black | 38 (6%) | 24 (6%) | 12 (7%) | 4 (6%) | 4 (6%) |
| Native American/Alaskan | 6 (1%) | 3 (1%) | 3 (2%) | 1 (2%) | 1 (2%) |
| Pacific Islander | 4 (1%) | 2 (1%) | 2 (1%) | 0 | 0 |
| White | 497 (83%) | 357 (86%) | 156 (85%) | 49 (88%) | 54 (87%) |
| Unknown | 24 (4%) | 18 (4%) | 8 (4%) | 2 (3%) | 2 (3%) |

WBC, white blood cell count; DA, daunorubicin and cytarabine; GO, gemtuzumab ozogamicin; CR, complete remission; MRD, measurable residual disease; NGS, next generation sequencing; DS, duplex sequencing

### Supplementary Table 3. Variants detected at diagnosis and remission.

| Patient | Gene | Transcript ID | HGVSc | HGVSp | Chr | Position | Reference | Alternate | Consequence | Diagnostic VAF | Remission VAF | VAF log fold change |
|---|---|---|---|---|---|---|---|---|---|---|---|---|
| SAASZM | CEBPA | NM_001287424.1 | c.494_495dupGC | p.Tyr166fs | chr19 | 33302024 | A | AGC | frameshift | 0.343 | N/A | N/A |
| SAASZM | NPM1 | NM_002520.6 | c.860_863dupTCTG | p.Trp288fs | chr5 | 171410539 | C | CTCTG | frameshift | 0.355 | 0.00010 | -3.54 |
| SAASZM | TET2 | NM_001127208.2 | c.715dupT | p.Ser239fs | chr4 | 105234654 | G | GT | frameshift | 0.340 | 0.01634 | -1.32 |
| SAASZM | TET2 | NM_001127208.2 | c.5440G>T | p.Gly1814* | chr4 | 105275950 | G | T | stop gained | 0.347 | 0.01643 | -1.32 |
| SAATEK | NRAS | NM_002524.4 | c.183A>C | p.Gln61His | chr1 | 114713907 | T | G | missense | 0.286 | N/A | N/A |
| SAATEK | NRAS | NM_002524.4 | c.181C>A | p.Gln61Lys | chr1 | 114713909 | G | T | missense | 0.114 | N/A | N/A |
| SAATEK | NRAS | NM_002524.4 | c.38G>A | p.Gly13Asp | chr1 | 114716123 | C | T | missense | 0.108 | N/A | N/A |
| SAATEK | RUNX1 | NM_001754.4 | c.1267C>T | p.Arg423Cys | chr21 | 34792311 | G | A | missense | 0.098 | N/A | N/A |
| SAATFD | FLT3 | NM_004119.2 | c.1745_1831dupCCGGCTCCTCAGATAATGAGTACTTCTACGTTGATTTCAGAGAATATGAATATGATCTCAAATGGGAGTTTCCAAGAGAAAATTTAG | p.Leu610_Glu611insAlaGlySerSerAspAsnGluTyrPheTyrValAspPheArgGluTyrGluTyrAspLeuLysTrpGluPheProArgGluAsnLeu | chr13 | 28034087 | T | TCTAAATTTTCTCTTGGAAACTCCCATTTGAGATCATATTCATATTCTCTGAAATCAACGTAGAAGTACTCATTATCTGAGGAGCCGG | ITD | 0.467 | N/A | N/A |
| SAATFD | FLT3 | NM_004119.2 | c.1831_1832insCCGGCTCCTCATATAATGAGTACTTCTACGTTGATTTCAGAGAATATGAATATGATCTCAAATGGGAGTTTCCAAGAGAAAATTTAG | p.Leu610_Glu611insAlaGlySerSerTyrAsnGluTyrPheTyrValAspPheArgGluTyrGluTyrAspLeuLysTrpGluPheProArgGluAsnLeu | chr13 | 28034087 | T | TCTAAATTTTCTCTTGGAAACTCCCATTTGAGATCATATTCATATTCTCTGAAATCAACGTAGAAGTACTCATTATATGAGGAGCCGG | ITD | 0.441 | N/A | N/A |
| SAATFD | IDH2 | NM_002168.3 | c.419G>A | p.Arg140Gln | chr15 | 90088702 | C | T | missense | 0.462 | 0.00976 | -1.67 |
| SAATFT | CBL | NM_005188.3 | c.1380_1382delTGA | p.Asp460del | chr11 | 119278648 | TATG | T | deletion | 0.054 | N/A | N/A |
| SAATFT | CEBPA | NM_001287424.1 | c.560C>G | p.Ala187Gly | chr19 | 33301960 | G | C | missense | 0.110 | N/A | N/A |
| SAATGC | DNMT3A | NM_022552.4 | c.2645G>A | p.Arg882His | chr2 | 25234373 | C | T | missense | 0.476 | 0.41760 | -0.06 |
| SAATGC | NPM1 | NM_002520.6 | c.860_863dupTCTG | p.Trp288fs | chr5 | 171410539 | C | CTCTG | frameshift | 0.392 | N/A | N/A |
| SAATGC | NRAS | NM_002524.4 | c.38G>A | p.Gly13Asp | chr1 | 114716123 | C | T | missense | 0.469 | N/A | N/A |
| SAATGG | CEBPA | NM_001287424.1 | c.1026_1031delCGTGGA | p.Asn342_Glu344delinsLys | chr19 | 33301494 | CTCCACG | C | deletion | 0.424 | N/A | N/A |
| SAATGG | CEBPA | NM_001287424.1 | c.172_173dupCC | p.His59fs | chr19 | 33302346 | C | CGG | frameshift | 0.386 | N/A | N/A |
| SAATGG | GATA2 | NM_032638.4 | c.961C>T | p.Leu321Phe | chr3 | 128483916 | G | A | missense | 0.423 | 0.00023 | -3.26 |
| SAATGG | NRAS | NM_002524.4 | c.38G>A | p.Gly13Asp | chr1 | 114716123 | C | T | missense | 0.235 | N/A | N/A |
| SAATHG | GATA2 | NM_032638.4 | c.1163_1165delTGA | p.Met388del | chr3 | 128481299 | TTCA | T | deletion | N/A | 0.15060 | N/A |
| SAATHG | IDH2 | NM_002168.3 | c.419G>A | p.Arg140Gln | chr15 | 90088702 | C | T | missense | 0.389 | 0.00987 | -1.60 |
| SAATHI | DNMT3A | NM_022552.4 | c.2644C>T | p.Arg882Cys | chr2 | 25234374 | G | A | missense | 0.411 | 0.27465 | -0.18 |
| SAATHI | FLT3 | NM_004119.2 | c.1751_1780dupCCTCAGATAATGAGTACTTCTACGTTGATT | p.Ser584_Asp593dup | chr13 | 28034138 | A | AAATCAACGTAGAAGTACTCATTATCTGAGG | ITD | 0.017 | 0.00052 | -1.50 |
| SAATHI | FLT3 | NM_004119.2 | c.1812_1813insAATGAGTACTTCTACGTTGATTTCAGAGAATATGAATATGATCTCG | p.Phe605fs | chr13 | 28034106 | A | ACGAGATCATATTCATATTCTCTGAAATCAACGTAGAAGTACTCATT | frameshift | N/A | 0.00262 | N/A |
| SAATHI | FLT3 | NM_004119.2 | c.1721_1804dupGCCAGCTACAGATGGTACAGGTGACCGGCTCCTCAGATAATGAGTACTTCTACGTTGATTTCAGAGAATATGAATATGATCTCA | p.Ser574_Leu601dup | chr13 | 28034114 | T | TTGAGATCATATTCATATTCTCTGAAATCAACGTAGAAGTACTCATTATCTGAGGAGCCGGTCACCTGTACCATCTGTAGCTGGC | ITD | N/A | 0.00396 | N/A |
| SAATHI | FLT3 | NM_004119.2 | c.1793_1794insCTACGTTGATTTCAGAGAATATGAATATGATCTCAAATGGGAGAATGAGTACTTCTACGTTGATTTCAGAGAATATGA | p.Tyr597_Glu598insAspTyrValAspPheArgGluTyrGluTyrAspLeuLysTrpGluAsnGluTyrPheTyrValAspPheArgGluTyr | chr13 | 28034125 | T | TTCATATTCTCTGAAATCAACGTAGAAGTACTCATTCTCCCATTTGAGATCATATTCATATTCTCTGAAATCAACGTAG | ITD | N/A | 0.00083 | N/A |
| SAATHI | NPM1 | NM_002520.6 | c.860_863dupTCTG | p.Trp288fs | chr5 | 171410539 | C | CTCTG | frameshift | 0.306 | 0.00437 | -1.85 |
| SAATHJ | EZH2 | NM_004456.4 | c.2232dupC | p.Glu745fs | chr7 | 148807669 | C | CG | frameshift | 0.669 | N/A | N/A |
| SAATHK | FLT3 | NM_004119.2 | c.1717_1794dupGAAAGCCAGCTACAGATGGTACAGGTGACCGGCTCCTCAGATAATGAGTACTTCTACGTTGATTTCAGAGAATATGAA | p.Glu598_Tyr599insGluSerGlnLeuGlnMetValGlnValThrGlySerSerAspAsnGluTyrPheTyrValAspPheArgGluTyrGlu | chr13 | 28034124 | A | ATTCATATTCTCTGAAATCAACGTAGAAGTACTCATTATCTGAGGAGCCGGTCACCTGTACCATCTGTAGCTGGCTTTC | ITD | 0.266 | N/A | N/A |
| SAATHK | FLT3 | NM_004119.2 | c.1794_1795insGAAAGCCAGCTACAGATGGTACAGGTTACCGGCTCCTCAGATAATGAGTACTTCTACGTTGATTTCAGAGAATATGAA | p.Glu598_Tyr599insGluSerGlnLeuGlnMetValGlnValThrGlySerSerAspAsnGluTyrPheTyrValAspPheArgGluTyrGlu | chr13 | 28034124 | A | ATTCATATTCTCTGAAATCAACGTAGAAGTACTCATTATCTGAGGAGCCGGTAACCTGTACCATCTGTAGCTGGCTTTC | ITD | 0.243 | N/A | N/A |
| SAATHK | FLT3 | NM_004119.2 | c.1794_1795insGAAAGCCAGCTACAGATGGTACAGGTGACCGGCTCCTCAGATAATGAGTACTTCTACGTTGATTTCAGAGAATATGATGAGTACTTCTACGTTGATTTCAGAGAATATGAA | p.Glu598_Tyr599insGluSerGlnLeuGlnMetValGlnValThrGlySerSerAspAsnGluTyrPheTyrValAspPheArgGluTyrAspGluTyrPheTyrValAspPheArgGluTyrGlu | chr13 | 28034124 | A | ATTCATATTCTCTGAAATCAACGTAGAAGTACTCATCATATTCTCTGAAATCAACGTAGAAGTACTCATTATCTGAGGAGCCGGTCACCTGTACCATCTGTAGCTGGCTTTC | ITD | 0.170 | N/A | N/A |
| SAATHK | FLT3 | NM_004119.2 | c.1761_1793dupTGAGTACTTCTACGTTGATTTCAGAGAATATGA | p.Tyr597_Glu598insAspGluTyrPheTyrValAspPheArgGluTyr | chr13 | 28034125 | T | TTCATATTCTCTGAAATCAACGTAGAAGTACTCA | ITD | 0.114 | N/A | N/A |
| SAATHK | IDH1 | NM_005896.3 | c.395G>A | p.Arg132His | chr2 | 208248388 | C | T | missense | 0.343 | N/A | N/A |
| SAATHK | NPM1 | NM_002520.6 | c.860_863dupTCTG | p.Trp288fs | chr5 | 171410539 | C | CTCTG | frameshift | 0.314 | N/A | N/A |
| SAATHK | PTPN11 | NM_002834.3 | c.181G>T | p.Asp61Tyr | chr12 | 112450361 | G | T | missense | 0.217 | N/A | N/A |
| SAATHK | RAD21 | NM_006265.2 | c.688+1G>T | | chr8 | 116857266 | C | A | splice donor | 0.348 | N/A | N/A |
| SAATHK | WT1 | NM_024426.4 | c.1390G>A | p.Asp464Asn | chr11 | 32392014 | C | T | missense | 0.477 | N/A | N/A |
| SAATHT | NRAS | NM_002524.4 | c.34G>T | p.Gly12Cys | chr1 | 114716127 | C | A | missense | 0.221 | N/A | N/A |
| SAATHW | FLT3 | NM_004119.2 | c.1796_1813dupATGATCTCAAATGGGAGT | p.Tyr599_Glu604dup | chr13 | 28034105 | A | AACTCCCATTTGAGATCAT | ITD | 0.197 | N/A | N/A |
| SAATHW | FLT3 | NM_004119.2 | c.1745_1786dupCCGGCTCCTCAGATAATGAGTACTTCTACGTTGATTTCAGAG | p.Arg595_Glu596insAlaGlySerSerAspAsnGluTyrPheTyrValAspPheArg | chr13 | 28034132 | T | TCTCTGAAATCAACGTAGAAGTACTCATTATCTGAGGAGCCGG | ITD | 0.163 | N/A | N/A |
| SAATHW | IDH2 | NM_002168.3 | c.418C>T | p.Arg140Trp | chr15 | 90088703 | G | A | missense | 0.432 | N/A | N/A |
| SAATHW | NPM1 | NM_002520.6 | c.860_863dupTCTG | p.Trp288fs | chr5 | 171410539 | C | CTCTG | frameshift | 0.014 | N/A | N/A |
| SAATHW | NPM1 | NM_002520.6 | c.863_864insCATG | p.Trp288fs | chr5 | 171410541 | C | CTGCA | frameshift | 0.325 | N/A | N/A |
| SAATHW | WT1 | NM_024426.4 | c.1390G>A | p.Asp464Asn | chr11 | 32392014 | C | T | missense | 0.537 | N/A | N/A |
| SAATJA | KIT | NM_000222.2 | c.1254_1255insTTCCTT | p.Tyr418_Asp419insPheLeu | chr4 | 54723606 | C | CTTCCTT | insertion | 0.439 | 0.00031 | -3.15 |
| SAATJA | WT1 | NM_024426.4 | c.1138_1142dupCGGTC | p.Ala382fs | chr11 | 32396363 | C | CGACCG | frameshift | 0.492 | 0.00038 | -3.11 |
| SAATKV | IDH2 | NM_002168.3 | c.419G>A | p.Arg140Gln | chr15 | 90088702 | C | T | missense | 0.410 | 0.00062 | -2.82 |
| SAATKV | NPM1 | NM_002520.6 | c.860_863dupTCTG | p.Trp288fs | chr5 | 171410539 | C | CTCTG | frameshift | 0.310 | N/A | N/A |
| SAATKY | KRAS | NM_004985.4 | c.183A>C | p.Gln61His | chr12 | 25227341 | T | G | missense | 0.380 | N/A | N/A |
| SAATKY | NPM1 | NM_002520.6 | c.863_864insCATG | p.Trp288fs | chr5 | 171410541 | C | CTGCA | frameshift | 0.320 | N/A | N/A |
| SAATKY | RAD21 | NM_006265.2 | c.1193delC | p.Pro398fs | chr8 | 116852677 | TG | T | frameshift | 0.058 | N/A | N/A |
| SAATKY | RAD21 | NM_006265.2 | c.1191_1192delACinsG | p.Pro398fs | chr8 | 116852679 | GT | C | frameshift | 0.232 | N/A | N/A |
| SAATLN | CBL | NM_005188.3 | c.1380_1382delTGA | p.Asp460del | chr11 | 119278648 | TATG | T | deletion | 0.053 | N/A | N/A |
| SAATLN | FLT3 | NM_004119.2 | c.2503G>T | p.Asp835Tyr | chr13 | 28018505 | C | A | missense | 0.128 | N/A | N/A |
| SAATNC | NRAS | NM_002524.4 | c.34G>T | p.Gly12Cys | chr1 | 114716127 | C | A | missense | 0.313 | 0.00025 | -3.10 |
| SAATNC | WT1 | NM_024426.4 | c.1142C>A | p.Ser381* | chr11 | 32396364 | G | T | stop gained | 0.369 | 0.00110 | -2.52 |

| Patient | Gene | Transcript ID | HGVSc | HGVSp | Chr | Position | Reference | Alternate | Consequence | Diagnostic VAF | Remission VAF | VAF log fold change |
|---|---|---|---|---|---|---|---|---|---|---|---|---|
| SAATND | CBL | NM_005188.3 | c.1096-25_1228-103del | | chr11 | 119278405 | ATAGTCTTTTAATTTTTTTTAATCAAAGGAACAATATGAATTATACTGTGAGATGGGCTCCACATTCCAACTATGTAAAATATGTGCTGAAAATGATAAGGATGTAAAGATTGAGCCCTGTGGACACCTCATGTGCACATCCTGTCTTACATCCTGGCAGGTACGGATCTAAACAGCGACTTTTTTCAGCTATGTAATAACCTTGGAAAATTCGGTATTATATAGCCTTTACTGATACAAGGGGTGGCCTGGCTTTTGGGGTTAGGTT | A | exon loss | 0.466 | N/A | N/A |
| SAATND | CBL | NM_005188.3 | c.1096-8_1228-86del | | chr11 | 119278416 | ATTTTTTTTAATCAAAGGAACAATATGAATTATACTGTGAGATGGGCTCCACATTCCAACTATGTAAAATATGTGCTGAAAATGATAAGGATGTAAAGATTGAGCCCTGTGGACACCTCATGTGCACATCCTGTCTTACATCCTGGCAGGTACGGATCTAAACAGCGACTTTTTTCAGCTATGTAATAACCTTGGAAAATTCGGTATTATATAGCCTTTACTGATACAAGGGGTGGCCTGGCTTTTGGGGTTAGGTTTAAACTTTTAC | A | exon loss | 0.130 | N/A | N/A |
| SAATRF | CEBPA | NM_001287424.1 | c.1039_1041dupCAG | p.Gln347dup | chr19 | 33301478 | T | TCTG | insertion | 0.311 | N/A | N/A |
| SAATRF | CEBPA | NM_001287424.1 | c.305_308delACAT | p.Tyr102fs | chr19 | 33302215 | GATGT | G | frameshift | 0.327 | N/A | N/A |
| SAATRF | RUNX1 | NM_001754.4 | c.1268G>A | p.Arg423His | chr21 | 34792310 | C | T | missense | 0.057 | N/A | N/A |
| SAATRF | WT1 | NM_024426.4 | c.1224dupA | p.Gln409fs | chr11 | 32396281 | G | GT | frameshift | 0.305 | N/A | N/A |
| SAATTF | FLT3 | NM_004119.2 | c.1719_1778dupAAGCCAGCTACAGATGGTACAGGTGACCGGCTCCTCAGATAATGAGTACTTCTACGTTGA | p.Glu573_Val592dup | chr13 | 28034140 | A | ATCAACGTAGAAGTACTCATTATCTGAGGAGCCGGTCACCTGTACCATCTGTAGCTGGCTT | ITD | 0.419 | N/A | N/A |
| SAATTF | NPM1 | NM_002520.6 | c.860_863dupTCTG | p.Trp288fs | chr5 | 171410539 | C | CTCTG | frameshift | 0.395 | N/A | N/A |
| SAATTF | TET2 | NM_001127208.2 | c.3629T>C | p.Leu1210Pro | chr4 | 105243604 | T | C | missense | 0.499 | N/A | N/A |
| SAATTF | TET2 | NM_001127208.2 | c.5541G>A | p.Trp1847* | chr4 | 105276051 | G | A | stop gained | 0.466 | N/A | N/A |
| SAATTL | FLT3 | NM_004119.2 | c.2503G>C | p.Asp835His | chr13 | 28018505 | C | G | missense | 0.326 | N/A | N/A |
| SAATTL | STAG2 | NM_001282418.1 | c.819+2_819+4delTAAinsAGAAGGGGC | | chrX | 124047509 | TAA | AGAAGGGGC | splice donor | 0.180 | N/A | N/A |
| SAATTL | STAG2 | NM_001282418.1 | c.819+6_819+8delCTTinsTTG | | chrX | 124047513 | CTT | TTG | splice region | 0.276 | N/A | N/A |
| SAATTL | WT1 | NM_024426.4 | c.1385G>C | p.Arg462Pro | chr11 | 32392019 | C | G | missense | 0.382 | N/A | N/A |
| SAATTR | PHF6 | NM_032458.2 | c.799_800insGA | p.Thr267fs | chrX | 134415084 | A | AAG | frameshift | 0.079 | N/A | N/A |
| SAATTR | WT1 | NM_024426.4 | c.1385G>A | p.Arg462Gln | chr11 | 32392019 | C | T | missense | 0.347 | 0.00445 | -1.89 |
| SAATVB | DNMT3A | NM_022552.4 | c.2644C>A | p.Arg882Ser | chr2 | 25234374 | G | T | missense | 0.372 | 0.00630 | -1.77 |
| SAATVB | NPM1 | NM_002520.6 | c.860_863dupTCTG | p.Trp288fs | chr5 | 171410539 | C | CTCTG | frameshift | 0.314 | N/A | N/A |
| SAATVX | FLT3 | NM_004119.2 | c.2503G>T | p.Asp835Tyr | chr13 | 28018505 | C | A | missense | 0.505 | 0.00017 | -3.48 |
| SAATWX | DNMT3A | NM_022552.4 | c.2644C>T | p.Arg882Cys | chr2 | 25234374 | G | A | missense | 0.456 | 0.00183 | -2.40 |
| SAATWX | KRAS | NM_004985.4 | c.164_181dupTTCTCGACACAGCAGGTC | p.Gly60_Gln61insLeuLeuAspThrAlaGly | chr12 | 25227342 | T | TGACCTGCTGTGTCGAGAA | insertion | 0.295 | N/A | N/A |
| SAATWX | KRAS | NM_004985.4 | c.175_176insTAGGTCTTCTCGACACAG | p.Thr58_Ala59insValGlyLeuLeuAspThr | chr12 | 25227348 | G | GCTGTGTCGAGAAGACCTA | insertion | 0.082 | N/A | N/A |
| SAATWX | NPM1 | NM_002520.6 | c.860_863dupTCTG | p.Trp288fs | chr5 | 171410539 | C | CTCTG | frameshift | 0.293 | 0.00006 | -3.70 |
| SAATWX | TET2 | NM_001127208.2 | c.1659delG | p.Pro555fs | chr4 | 105235601 | TG | T | frameshift | 0.462 | 0.00238 | -2.29 |
| SAATYG | EZH2 | NM_004456.4 | c.2226_2227insAAT | p.Val742_Gly743insAsn | chr7 | 148807675 | C | CATT | insertion | 0.815 | 0.00009 | -3.96 |
| SAATYG | FLT3 | NM_004119.2 | c.2504A>T | p.Asp835Val | chr13 | 28018504 | T | A | missense | 0.182 | N/A | N/A |
| SAATYG | FLT3 | NM_004119.2 | c.1821_1822insTTCCCTAATGAATATGATCTCAAATGGGAGTTTCCAAGA | p.Arg607_Glu608insPheProAsnGluTyrAspLeuLysTrpGluPheProArg | chr13 | 28034097 | C | CTCTTGGAAACTCCCATTTGAGATCATATTCATTAGGGAA | ITD | 0.207 | N/A | N/A |
| SAATYG | TP53 | NM_000546.5 | c.711G>C | p.Met237Ile | chr17 | 7674252 | C | G | missense | N/A | 0.01087 | N/A |
| SAATYG | U2AF1 | NM_006758.2.2 | c.101C>A | p.Ser34Tyr | chr21 | 43104346 | G | T | missense | 0.489 | 0.00543 | -1.95 |
| SAATYL | CBL | NM_005188.3 | c.1259G>A | p.Arg420Gln | chr11 | 119278541 | G | A | missense | N/A | 0.00155 | N/A |
| SAATYL | DNMT3A | NM_022552.4 | c.2645G>A | p.Arg882His | chr2 | 25234373 | C | T | missense | 0.233 | 0.00014 | -3.21 |
| SAATYL | NPM1 | NM_002520.6 | c.860_863dupTCTG | p.Trp288fs | chr5 | 171410539 | C | CTCTG | frameshift | 0.199 | N/A | N/A |
| SAATZW | TP53 | NM_000546.5 | c.844C>T | p.Arg282Trp | chr17 | 7673776 | G | A | missense | 0.647 | 0.13124 | -0.69 |
| SAAUBG | EZH2 | NM_004456.4 | c.2187dupT | p.Asp730fs | chr7 | 148809078 | C | CA | frameshift | 0.380 | 0.00011 | -3.55 |
| SAAUBG | KIT | NM_000222.2 | c.2447A>T | p.Asp816Val | chr4 | 54733155 | A | T | missense | 0.058 | 0.00007 | -2.92 |
| SAAUBG | RAD21 | NM_006265.2 | c.2T>C | p.Met1? | chr8 | 116866728 | A | G | start lost | 0.055 | N/A | N/A |
| SAAUBK | NRAS | NM_002524.4 | c.34G>A | p.Gly12Ser | chr1 | 114716127 | C | T | missense | 0.195 | N/A | N/A |
| SAAUBW | FLT3 | NM_004119.2 | c.1770_1829dupCTACGTTGATTTCAGAGAATATGAATATGATCTCAAATGGGAGTTTCCAAGAGAAAATTT | p.Phe590_Asn609dup | chr13 | 28034089 | T | TAAATTTTCTCTTGGAAACTCCCATTTGAGATCATATTCATATTCTCTGAAATCAACGTAG | ITD | 0.399 | 0.00034 | -3.07 |
| SAAUBW | NPM1 | NM_002520.6 | c.860_863dupTCTG | p.Trp288fs | chr5 | 171410539 | C | CTCTG | frameshift | 0.387 | 0.00035 | -3.05 |
| SAAUBW | SMC1A | NM_006306.3 | c.1880G>T | p.Arg627Leu | chrX | 53405524 | C | A | missense | N/A | 0.00126 | N/A |
| SAAUBW | TET2 | NM_001127208.2 | c.4097G>A | p.Arg1366His | chr4 | 105269662 | G | A | missense | 0.502 | 0.02946 | -1.23 |
| SAAUBZ | IDH2 | NM_002168.3 | c.418C>T | p.Arg140Trp | chr15 | 90088703 | G | A | missense | 0.404 | 0.00012 | -3.54 |
| SAAUCC | FLT3 | NM_004119.2 | c.1786_1866dupGAATATGAATATGATCTCAAATGGGAGTTTCCAAGAGAAAATTTAGAGTTTGGTAAGAATGGAATGTGCCAAATGTTTCTGCAGCATTTCTTTTCCATTGGAAAATCTTTAAAATGCACGTACTCACCATTTGTCTTTGCAGGGAAGGTACTAGGATCAGGTGCTTTTGGA | p.Gly622_Lys623insGluTyrGluTyrAspLeuLysTrpGluPheProArgGluAsnLeuGluPheGlyLysAsnGlyMetCysGlnMetPheLeuGlnHisPhePheSerIleGlyLysSerLeuLysCysThrTyrSerProPheValPheAlaGlyLysValLeuGlySerGlyAlaPheGly | chr13 | 28033962 | T | TTCCAAAAGCACCTGATCCTAGTACCTTCCCTGCAAAGACAAATGGTGAGTACGTGCATTTTAAAGATTTTCCAATGGAAAAGAAATGCTGCAGAAACATTTGGCACATTCCATTCTTACCAAACTCTAAATTTTCTCTTGGAAACTCCCATTTGAGATCATATTCATATTC | ITD | 0.204 | 0.00044 | -2.66 |
| SAAUCC | FLT3 | NM_004119.2 | c.1773_1799dupCGTTGATTTCAGAGAATATGAATATGA | p.Asp600_Leu601insValAspPheArgGluTyrGluTyrAsp | chr13 | 28034119 | A | ATCATATTCATATTCTCTGAAATCAACG | ITD | 0.078 | 0.00025 | -2.49 |
| SAAUCC | NPM1 | NM_002520.6 | c.860_863dupTCTG | p.Trp288fs | chr5 | 171410539 | C | CTCTG | frameshift | 0.348 | 0.02134 | -1.21 |
| SAAUCC | NRAS | NM_002524.4 | c.34G>T | p.Gly12Cys | chr1 | 114716127 | C | A | missense | 0.300 | 0.00043 | -2.85 |
| SAAUCC | PTPN11 | NM_002834.3 | c.1530G>C | p.Gln510His | chr12 | 112489106 | G | C | missense | 0.059 | 0.01061 | -0.74 |
| SAAUCG | IDH1 | NM_005896.3 | c.395G>A | p.Arg132His | chr2 | 208248388 | C | T | missense | 0.089 | 0.00723 | -1.09 |
| SAAUCI | KIT | NM_000222.2 | c.2466T>G | p.Asn822Lys | chr4 | 54733174 | T | G | missense | 0.100 | 0.00007 | -3.18 |
| SAAUCI | KRAS | NM_004985.4 | c.38_39delGCinsAT | p.Gly13Asp | chr12 | 25245347 | GC | AT | missense | 0.118 | 0.00047 | -2.40 |
| SAAUCK | ASXL1 | NM_015338.5 | c.1774C>T | p.Gln592* | chr20 | 32434486 | C | T | stop gained | 0.125 | N/A | N/A |
| SAAUCL | KIT | NM_000222.2 | c.2447A>T | p.Asp816Val | chr4 | 54733155 | A | T | missense | 0.375 | 0.00048 | -2.89 |
| SAAUCP | DNMT3A | NM_022552.4 | c.2645G>A | p.Arg882His | chr2 | 25234373 | C | T | missense | 0.251 | 0.09516 | -0.42 |
| SAAUCP | NPM1 | NM_002520.6 | c.860_863dupTCTG | p.Trp288fs | chr5 | 171410539 | C | CTCTG | frameshift | 0.153 | 0.00064 | -2.38 |

| Patient | Gene | Transcript ID | HGVSc | HGVSp | Chr | Position | Reference | Alternate | Consequence | Diagnostic VAF | Remission VAF | VAF log fold change |
|---|---|---|---|---|---|---|---|---|---|---|---|---|
| SAAUCP | NRAS | NM_002524.4 | c.182A>G | p.Gln61Arg | chr1 | 114713908 | T | C | missense | 0.187 | 0.00043 | -2.64 |
| SAAUCP | NRAS | NM_002524.4 | c.35G>A | p.Gly12Asp | chr1 | 114716126 | C | T | missense | 0.117 | N/A | N/A |
| SAAUCP | SMC1A | NM_006306.3 | c.2141A>G | p.Tyr714Cys | chrX | 53405067 | T | C | missense | N/A | 0.00209 | N/A |
| SAAUCW | FLT3 | NM_004119.2 | c.1837+1_1837+2insAAGGGTTTTCAGAGAATATGAATATGATCTCAAATGGGAGTTTCCAAGAGAAAATTTAGAGTTTGG |  | chr13 | 28034080 | A | ACCAAACTCTAAATTTTCTCTTGGAAACTCCCATTTGAGATCATATTCATATTCTCTGAAAACCCTT | ITD | 0.421 | N/A | N/A |
| SAAUCW | PHF6 | NM_032458.2 | c.27dupA | p.Gly10fs | chrX | 134377638 | G | GA | frameshift | 0.836 | N/A | N/A |
| SAAUCW | RUNX1 | NM_001754.4 | c.1268_1276delGCTCGCCGCinsCACCCCG | p.Arg423fs | chr21 | 34792310 | GCGGCGAGC | CGGGGTG | frameshift | 0.460 | N/A | N/A |
| SAAUDH | DNMT3A | NM_022552.4 | c.2645G>A | p.Arg882His | chr2 | 25234373 | C | T | missense | 0.448 | 0.33074 | -0.13 |
| SAAUDH | NPM1 | NM_002520.6 | c.860_863dupTCTG | p.Trp288fs | chr5 | 171410539 | C | CTCTG | frameshift | 0.331 | 0.00005 | -3.81 |
| SAAUDH | PTPN11 | NM_002834.3 | c.1508G>T | p.Gly503Val | chr12 | 112489084 | G | T | missense | 0.425 | N/A | N/A |
| SAAUDW | DNMT3A | NM_022552.4 | c.1154delC | p.Pro385fs | chr2 | 25246745 | CG | C | frameshift | 0.408 | 0.00174 | -2.37 |
| SAAUDW | IDH1 | NM_005896.3 | c.395G>A | p.Arg132His | chr2 | 208248388 | C | T | missense | 0.443 | 0.00019 | -3.36 |
| SAAUDW | NPM1 | NM_002520.6 | c.863_864insCTTG | p.Trp288fs | chr5 | 171410541 | C | CTGCT | frameshift | 0.426 | 0.00018 | -3.37 |
| SAAUDW | PTPN11 | NM_002834.3 | c.179G>T | p.Gly60Val | chr12 | 112450359 | G | T | missense | 0.418 | 0.00029 | -3.16 |
| SAAUDW | RAD21 | NM_006265.2 | c.1660_1663delAGAA | p.Arg554fs | chr8 | 116848990 | CTTCT | C | frameshift | 0.395 | 0.00057 | -2.84 |
| SAAUED | RAD21 | NM_006265.2 | c.1818_1820delGCA | p.Gln607del | chr8 | 116847578 | TTGC | T | deletion | 0.415 | N/A | N/A |
| SAAUEE | FLT3 | NM_004119.2 | c.1780_1800dupTTCAGAGAATATGAATATGAT | p.Asp600_Leu601insPheArgGluTyrGluTyrAsp | chr13 | 28034118 | G | GATCATATTCATATTCTCTGAA | ITD | 0.290 | 0.00053 | -2.74 |
| SAAUEE | FLT3 | NM_004119.2 | c.1778_1779insCTTCAGAGAATATGAATATGA | p.Asp593_Phe594insPheArgGluTyrGluTyrAsp | chr13 | 28034140 | A | ATCATATTCATATTCTCTGAAG | ITD | 0.080 | N/A | N/A |
| SAAUEE | IDH2 | NM_002168.3 | c.419G>A | p.Arg140Gln | chr15 | 90088702 | C | T | missense | 0.436 | 0.02294 | -1.28 |
| SAAUEE | NPM1 | NM_002520.6 | c.860_863dupTCTG | p.Trp288fs | chr5 | 171410539 | C | CTCTG | frameshift | 0.398 | 0.01258 | -1.50 |
| SAAUEG | ASXL1 | NM_015338.5 | c.3031delG | p.Glu1011fs | chr20 | 32435742 | CG | C | frameshift | 0.075 | N/A | N/A |
| SAAUEG | PHF6 | NM_032458.2 | c.319delT | p.Cys107fs | chrX | 134393579 | T | <DEL> | frameshift | N/A | 0.02163 | N/A |
| SAAUEG | WT1 | NM_024426.4 | c.1369_1405delCAGCGAAAGTTCTCCCGGTCCGACCACCTGAAGACCCinsTATTTTTTTTTTTATGTTTCGGAGAACATGTAGATCT | p.GlnArgLysPheSerArgSerAspHisLeuLysThrHis457* | chr11 | 32392035 | GGGTCTTCAGGTGGTCGGACCGGGAGAACTTTCGCTG | AGATCTACATGTTCTCCGAAACATAAAAAAAAAAATA | stop gained | 0.052 | N/A | N/A |
| SAAUEL | CEBPA | NM_001287424.1 | c.1049_1050insCAC | p.Leu350_Glu351insThr | chr19 | 33301470 | C | CGTG | insertion | 0.423 | N/A | N/A |
| SAAUEL | CEBPA | NM_001287424.1 | c.440_441dupCC | p.Ala148fs | chr19 | 33302078 | C | CGG | frameshift | 0.358 | N/A | N/A |
| SAAUEL | FLT3 | NM_004119.2 | c.1730_1801dupAGATGGTACAGGTGACCGGCTCCTCAGATAATGAGTACTTCTACGTTGATTTCAGAGAATATGAATATGATC | p.Gln577_Asp600dup | chr13 | 28034117 | A | AGATCATATTCATATTCTCTGAAATCAACGTAGAAGTACTCATTATCTGAGGAGCCGGTCACCTGTACCATCT | ITD | 0.487 | N/A | N/A |
| SAAUEL | NRAS | NM_002524.4 | c.35G>A | p.Gly12Asp | chr1 | 114716126 | C | T | missense | 0.052 | N/A | N/A |
| SAAUEL | RAD21 | NM_006265.2 | c.193C>T | p.Arg65* | chr8 | 116863211 | G | A | stop gained | 0.426 | 0.00016 | -3.44 |
| SAAUEY | KIT | NM_000222.2 | c.1248_1262delGACTTACGACAGGCTinsCCGGTCTTA | p.Thr417_Leu421delinsArgSerTyr | chr4 | 54723614 | GACTTACGACAGGCT | CCGGTCTTA | indel | 0.373 | 0.00061 | -2.79 |
| SAAUEY | WT1 | NM_024426.4 | c.1107delAinsGG | p.Arg370fs | chr11 | 32396399 | T | CC | frameshift | 0.407 | 0.00028 | -3.17 |
| SAAUFB | FLT3 | NM_004119.2 | c.1788_1789insGTTAATGAGTACTTCTACGTTGATTTCAGAGAA | p.Glu596_Tyr597insValAsnGluTyrPheTyrValAspPheArgGlu | chr13 | 28034130 | A | ATTCTCTGAAATCAACGTAGAAGTACTCATTAAC | ITD | 0.459 | 0.00009 | -3.69 |
| SAAUFB | WT1 | NM_024426.4 | c.1141_1144dupTCGG | p.Ala382fs | chr11 | 32396361 | G | GCCGA | frameshift | 0.406 | 0.00006 | -3.86 |
| SAAUFE | CBL | NM_005188.3 | c.1137_1147delACTATGTAAAA | p.Gln379fs | chr11 | 119278216 | CAACTATGTAAA | C | frameshift | N/A | 0.00101 | N/A |
| SAAUFE | DNMT3A | NM_022552.4 | c.2645G>A | p.Arg882His | chr2 | 25234373 | C | T | missense | 0.455 | 0.26550 | -0.23 |
| SAAUFE | IDH1 | NM_005896.3 | c.394C>T | p.Arg132Cys | chr2 | 208248389 | G | A | missense | 0.548 | 0.08209 | -0.82 |
| SAAUFE | RUNX1 | NM_001754.4 | c.958C>T | p.Arg320* | chr21 | 34799310 | G | A | stop gained | N/A | 0.00156 | N/A |
| SAAUFF | FLT3 | NM_004119.2 | c.1786_1824dupGAATATGAATATGATCTCAAATGGGAGTTTCCAAGAGAA | p.Glu608_Asn609insGluTyrGluTyrAspLeuLysTrpGluPheProArgGlu | chr13 | 28034094 | T | TTTCTCTTGGAAACTCCCATTTGAGATCATATTCATATTC | ITD | 0.240 | 0.00016 | -3.17 |
| SAAUFN | NRAS | NM_002524.4 | c.35G>A | p.Gly12Asp | chr1 | 114716126 | C | T | missense | 0.421 | 0.00020 | -3.33 |
| SAAUFU | NRAS | NM_002524.4 | c.35G>A | p.Gly12Asp | chr1 | 114716126 | C | T | missense | 0.350 | 0.00098 | -2.55 |
| SAAUFU | GATA2 | NM_032638.4 | c.1203_1233delCAACAAGTCCAAGAAGAGCAAGAAAGGGGCGinsGGCA | p.Asn402_Gly410del | chr13 | 128481259 | CGCCCCTTTCTTGCTCTTCTTGGACTTGTTG | TGCC | indel | N/A | 0.01450 | N/A |
| SAAUFV | DNMT3A | NM_022552.4 | c.2204A>G | p.Tyr735Cys | chr2 | 25240420 | T | C | missense | 0.397 | 0.00444 | -1.95 |
| SAAUFV | DNMT3A | NM_022552.4 | c.976C>T | p.Arg326Cys | chr2 | 25247629 | G | A | missense | 0.369 | 0.16054 | -0.36 |
| SAAUFV | KIT | NM_000222.2 | c.2447A>T | p.Asp816Val | chr4 | 54733155 | A | T | missense | 0.355 | N/A | N/A |
| SAAUGM | FLT3 | NM_004119.2 | c.2503G>T | p.Asp835Tyr | chr13 | 28018505 | C | A | missense | 0.432 | N/A | N/A |
| SAAUHE | ASXL1 | NM_015338.5 | c.1900_1922delAGAGAGGCGGCCACCACTGCCAT | p.Glu635fs | chr20 | 32434622 | TCACCACTGCCATAGAGAGGCGGC | T | frameshift | 0.133 | N/A | N/A |
| SAAUHE | CBL | NM_005188.3 | c.1380_1382delTGA | p.Asp460del | chr11 | 119278648 | TATG | T | deletion | 0.052 | N/A | N/A |
| SAAUHE | CEBPA | NM_001287424.1 | c.560C>G | p.Ala187Gly | chr19 | 33301960 | G | C | missense | 0.087 | N/A | N/A |
| SAAUHE | KIT | NM_000222.2 | c.2466T>A | p.Asn822Lys | chr4 | 54733174 | T | A | missense | 0.182 | N/A | N/A |
| SAAUHE | KIT | NM_000222.2 | c.2466T>G | p.Asn822Lys | chr4 | 54733174 | T | G | missense | 0.160 | N/A | N/A |
| SAAUHH | DNMT3A | NM_022552.4 | c.2654G>A | p.Arg885Lys | chr2 | 25234364 | C | T | missense | 0.459 | 0.00023 | -3.31 |
| SAAUHH | FLT3 | NM_004119.2 | c.2523C>A | p.Asn841Lys | chr13 | 28018485 | G | T | missense | 0.145 | N/A | N/A |
| SAAUHH | NPM1 | NM_002520.6 | c.860_863dupTCTG | p.Trp288fs | chr5 | 171410539 | C | CTCTG | frameshift | 0.314 | 0.00019 | -3.21 |
| SAAUHH | PTPN11 | NM_002834.3 | c.182A>T | p.Asp61Val | chr12 | 112450362 | A | T | missense | 0.263 | N/A | N/A |
| SAAUHY | DNMT3A | NM_022552.4 | c.2645G>A | p.Arg882His | chr2 | 25234373 | C | T | missense | 0.535 | 0.06667 | -0.90 |
| SAAUHY | NPM1 | NM_002520.6 | c.860_863dupTCTG | p.Trp288fs | chr5 | 171410539 | C | CTCTG | frameshift | 0.368 | 0.01162 | -1.50 |
| SAAUHY | NRAS | NM_002524.4 | c.182A>G | p.Gln61Arg | chr1 | 114713908 | T | C | missense | 0.379 | 0.00074 | -2.71 |
| SAAUHY | SMC1A | NM_006306.3 | c.1468G>A | p.Asp490Asn | chrX | 53409139 | C | T | missense | 0.915 | 0.03895 | -1.37 |
| SAAUKF | KRAS | NM_004985.4 | c.35G>A | p.Gly12Asp | chr12 | 25245350 | C | T | missense | 0.025 | 0.00135 | -1.28 |
| SAAUKF | KRAS | NM_004985.4 | c.34G>A | p.Gly12Ser | chr12 | 25245351 | C | T | missense | 0.252 | 0.01180 | -1.33 |
| SAAUKF | TP53 | NM_000546.5 | c.499_500delCA | p.Gln167fs | chr17 | 7675113 | CTG | C | frameshift | 0.675 | 0.00986 | -1.84 |
| SAAVFE | DNMT3A | NM_022552.4 | c.2645G>A | p.Arg882His | chr2 | 25234373 | C | T | missense | 0.086 | 0.00008 | -3.01 |
| SAAVFE | FLT3 | NM_004119.2 | c.1837+3_1837+4insCCCTTCTCAGAGAATATGAATATGATCTCAAATGGGAGTTTCCAAGAGAAAATTTAGAGTTTGGTA |  | chr13 | 28034078 | T | TTACCAAACTCTAAATTTTCTCTTGGAAACTCCCATTTGAGATCATATTCATATTCTCTGAGAAGGG | ITD | 0.615 | 0.00011 | -3.74 |
| SAAVFE | FLT3 | NM_004119.2 | c.1837+3_1837+4insCCCTTCTCAGAGAATATGCATATGATCTCAAATGGGAGTTTCCAAGAGAAAATTTAGAGTTTGGTA |  | chr13 | 28034078 | T | TTACCAAACTCTAAATTTTCTCTTGGAAACTCCCATTTGAGATCATATGCATATTCTCTGAGAAGGG | ITD | 0.467 | N/A | N/A |
| SAAVFE | RUNX1 | NM_001754.4 | c.1267C>T | p.Arg423Cys | chr21 | 34792311 | G | A | missense | 0.158 | N/A | N/A |
| SAAVFE | STAG2 | NM_001282418.1 | c.3397C>T | p.Arg1133Trp | chrX | 124090694 | C | T | missense | 0.154 | N/A | N/A |
| SAAVHJ | RUNX1 | NM_001754.4 | c.501T>G | p.Ser167Arg | chr21 | 34880564 | A | C | missense | 0.450 | 0.01486 | -1.48 |
| SAAVID | DNMT3A | NM_022552.4 | c.2644C>T | p.Arg882Cys | chr2 | 25234374 | G | A | missense | 0.219 | 0.01658 | -1.12 |
| SAAVID | IDH2 | NM_002168.3 | c.419G>A | p.Arg140Gln | chr15 | 90088702 | C | T | missense | 0.164 | 0.01047 | -1.20 |
| SAAVID | RUNX1 | NM_001754.4 | c.485G>A | p.Arg162Lys | chr21 | 34880580 | C | T | missense | 0.211 | 0.00031 | -2.83 |
| SAAVJA | CEBPA | NM_001287424.1 | c.1040_1041insCCGCGACAAGGCCAAGCAGCGCAACGTGGAGACGCCGCA | p.Gln346_Gln347insHisArgAspLysAlaLysGlnArgAsnValGluThrPro | chr19 | 33301479 | C | CTGCGGCGTCTCCACGTTGCGCTGCTTGGCCTTGTCGCGG | insertion | 0.149 | N/A | N/A |
| SAAVJA | CEBPA | NM_001287424.1 | c.222delC | p.Ala75fs | chr19 | 33302298 | CG | C | frameshift | 0.262 | N/A | N/A |
| SAAVJA | GATA2 | NM_032638.4 | c.923G>C | p.Arg308Pro | chr3 | 128483954 | C | G | missense | 0.344 | 0.00010 | -3.53 |
| SAAVJP | DNMT3A | NM_022552.4 | c.2645G>A | p.Arg882His | chr2 | 25234373 | C | T | missense | 0.430 | 0.02000 | -1.33 |
| SAAVJP | FLT3 | NM_004119.2 | c.1779_1793dupTTTCAGAGAATATGA | p.Asp593_Tyr597dup | chr13 | 28034125 | T | TTCATATTCTCTGAAA | ITD | 0.415 | 0.00007 | -3.80 |
| SAAVJP | NPM1 | NM_002520.6 | c.860_863dupTCTG | p.Trp288fs | chr5 | 171410539 | C | CTCTG | frameshift | 0.294 | N/A | N/A |

**Supplementary Table 4. Flow cytometry MRD.**

| Patient | Flow MRD (%) |
|---|---|
| SAATHG | 0.28 |
| SAATLN | 0.002 |
| SAATTR | 0.6 |
| SAAUCW | 0.01 |
| SAAUEE | 2.3 |
| SAAUEG | 0.21 |
| SAAUFE | 6.2 |
| SAAUFU | 0.01 |
| SAAUKF | 0.2 |
| SAAVJP | 0.85 |

**Supplementary Table 5. Univariate cox regression analysis for clinical outcomes based on various duplex sequencing MRD definitions.**

| MRD Definition | Relapse | | Relapse-Free Survival | | Overall Survival | |
|---|---|---|---|---|---|---|
| | HR (95% CI) | p-value | HR (95% CI) | p-value | HR (95% CI) | p-value |
| DS - any RDV | 2.8 (0.8-9.4) | 0.11 | 1.6 (0.7-4) | 0.3 | 1.5 (0.6-4.2) | 0.39 |
| DS - RDV with VAF ≥0.1% | 4.7 (1.8-12.3) | 0.0016 | 3.8 (1.7-8.5) | 0.0014 | 3.2 (1.3-7.7) | 0.0088 |
| DS - RDV with VAF ≥0.1%, no DTA | 5.5 (2.3-13.3) | <0.001 | 4.6 (2.1-10) | <0.001 | 4.8 (2-11.1) | <0.001 |
| DS - RDV ≤2 $log^{10}$ reduction in VAF between diagnosis and remission | 3.3 (1.4-8.2) | 0.0086 | 2.6 (1.2-5.6) | 0.017 | 2.1 (0.9-4.8) | 0.089 |
| DS - RDV ≤2 $log^{10}$ reduction in VAF between diagnosis and remission, no DTA | 4.5 (1.9-11) | <0.001 | 4 (1.8-8.7) | <0.001 | 4.3 (1.8-10.1) | <0.001 |
| DS - VAF ≥0.1% or ≥0.01% for *NPM1/FLT3*-ITD, no DTA, agnostic to diagnosis | 8.7 (2.9-26.1) | <0.001 | 4.8 (2.1-11.1) | <0.001 | 5.4 (2.1-13.8) | <0.001 |

MRD, measurable residual disease; HR, hazard ratio; CI, confidence interval; DS, duplex sequencing; RDV, residual diagnostic variant; VAF, variant allele frequency; DTA, *DNMT3A TET2 ASXL1*
